# AutoML-Multiverse: An Instability-Aware Framework for Quantifying Analytic Variability in Alzheimer’s Disease Machine-Learning Studies

**DOI:** 10.64898/2026.03.13.26347938

**Authors:** Maitrei Kohli, Gonzalo Castro Leal, Douglas Wyllie, Neil Oxtoby, Robert Leech, Philip Weston, James Cole, the Alzheimer’s Disease Neuroimaging Initiative

## Abstract

Machine-learning (ML) models for Alzheimer’s disease (AD) frequently yield divergent conclusions, raising concerns about robustness, reproducibility, and interpretability. This instability is partially driven by researcher biases and analytic variability. Coupled with the clinical heterogeneity, mixed pathologies, and cohort differences in AD research, these issues limit the reliability and validity of conclusions from individual models.

We introduce AutoML-Multiverse, an instability-aware framework characterising how analytic choices influence ML-based conclusions. The AutoML-Multiverse explores a large space of ∼20,000 analysis pipelines and by retaining the full distribution of pipelines, enables direct examination of analytic variability. We evaluate this framework across 20 classification tasks in two independent cohorts studying Alzheimer’s disease progression (ADNI, N≤1,930; NACC, N≤1,057), using multiple data modalities: neuroimaging, clinical/cognitive and fluid biomarkers.

AutoML-Multiverse performance was equal to or better than non-automated models across all tasks. For example, stable versus progressive mild cognitive impairment (MCI) classification accuracy was 0.68±0.06 (ADNI) and 0.63±0.08 (NACC), while AD versus cognitively normal (CN) classification reached 0.97±0.01 (ADNI). Crucially, each modality’s utility was task- and cohort-dependent: clinical measures dominated diagnostic tasks, whereas imaging better predicted progression, with modality preferences often switching between cohorts, highlighting limited generalisability of single-cohort results.

Using the AutoML-Multiverse, we obtained strong classification performance without pre-specifying key model design choices. By reducing analysis-driven variability and explicitly characterising uncertainty, instability-aware evaluation can support the development of more robust and clinically applicable prediction models in AD research.

**Highlights:**

- AutoML-Multiverse systematically quantifies analytic instability in clinical ML.
- Analysis of ∼20,000 pipelines across ADNI and NACC cohorts.
- Pipeline choices substantially alter model rankings and biomarker importance.
- Cross-cohort variability highlights risks of single-dataset studies.
- Instability-aware evaluation improves robustness of AI-driven research.

## 1. Introduction

Machine learning (ML) approaches are increasingly used in Alzheimer’s disease (AD) research to predict diagnosis, prognosis, and disease staging using neuroimaging, clinical, and biomarker data. Applications include distinguishing cognitively normal (CN) individuals from those with mild cognitive impairment (MCI) or AD, predicting progression from stable to progressive MCI, and integrating multimodal information to improve predictive performance(1)(2)(3). Despite rapid methodological advances and steadily improving performance metrics, the reliability and robustness of scientific and clinical conclusions drawn from such models remain a critical concern for clinical translation(4).

Even when modelling objectives are clearly defined and analyses adhere to established standards (e.g., standard preprocessing and validation procedures)(5), results from ML-based AD studies can vary substantially depending on analytic and modelling decisions (4)(6)(7).In a landmark many-analysts study (not specific to Alzheimer’s disease), Botvinik-Nezer et al. demonstrated that 70 independent research teams analysing the same neuroimaging dataset, while addressing the same scientific question, arrived at markedly different conclusions primarily driven by differences in analytical choices (8). Subsequent studies have reinforced that such analytic flexibility is not an anomaly, but is often inherent in complex data analyses, with important implications for reproducibility and scientific inference (9)(10)(11).

In the context of AD research, this sensitivity to analytical decisions is particularly consequential. ML studies often involve multiple, equally valid choices related to model and pipeline specification, data sampling and resampling strategies, selection of data modalities, and cohort composition, each of which can substantially influence performance and downstream conclusions(4)(6). As a result, different yet reasonable analytical pipelines may yield divergent estimates of performance, inconsistent assessments of modality importance, or even conflicting conclusions about disease-related patterns.

This analytical sensitivity is further amplified by the biological and clinical heterogeneity of AD itself. AD represents a clinical and pathological spectrum rather than a single uniform entity, encompassing mixed pathologies, variable trajectories of amyloid and tau accumulation, and substantial inter-individual differences in cognitive resilience and comorbidity(12)(4). In this setting, a single “optimal” modelling pipeline may preferentially capture signals associated with a subset of disease trajectories or pathological profiles, while obscuring others, thereby limiting the generalisability and clinical interpretability of the resulting conclusions.

The growing recognition of analytic flexibility and researcher induced biases has motivated the development of automated approaches aimed at systematically navigating complex modelling choices while reducing reliance on ad hoc analyst decisions. Automated machine learning (AutoML) frameworks were introduced to formalise and automate key stages of the ML pipeline, including model selection, hyperparameter optimisation, feature preprocessing, and, in some cases, feature selection and model ensembling (13)(14)(15). By algorithmically exploring large spaces of candidate pipelines under predefined constraints, AutoML seeks to improve efficiency and reproducibility while lowering the technical barrier to applying ML methods in applied biomedical domains(16)(17).

In applied biomedical and neuroimaging research, AutoML has predominantly been adopted as a performance-optimisation tool, with success typically evaluated using predictive accuracy or area under the receiver operating characteristic curve. Numerous studies report that AutoML pipelines outperform manually selected ML models or achieve performance comparable to deep learning approaches, particularly in settings involving structured clinical and imaging data (18)(19)(20)(21). Consequently, most AutoML studies emphasise the identification and reporting of a single best-performing pipeline, often accompanied by an ensemble constructed from top-ranked candidates(22)(18)(19)(23).

Typically, autoML frameworks are deployed under implicit assumptions, including the existence of a single optimal pipeline for a given task, the expectation that increased search yields stable performance estimates, and the interpretation of the final selected model as a reliable basis for inference regarding feature or modality importance. However, Dafflon et al. (2022) formalised analytical flexibility using a multiverse framework (5), defining decision points and systematically combining them to generate alternative pipelines. This approach directly quantifies how conclusions vary across the decision space and shows that resulting variability can arise from the structure of the analytic space itself, even when analyses are fully specified and reproducible.

Despite the popularity of ML in AD research, relatively little work has examined how sensitive ML-derived conclusions are to variation across modelling pipelines, data sampling strategies, data modalities, and cohort characteristics. As a result, analytical variability is frequently overlooked, and just the single best-performing model reported. This leaves open questions regarding the robustness and generalisability of the resulting conclusions, particularly in clinical contexts where stable and reproducible evidence is essential.

Building on multiverse analyses that characterise variability across analytical decision spaces (5) and recent work on large-scale exploration of modelling pipelines(24), we introduce the autoML-Multiverse as an instability-aware evaluation framework. By retaining and aggregating information across multiple, complementary pipelines, instead of a winner-takes-all focus on the single best-performing model, our framework enables assessment of robustness and consistency across prediction tasks. The autoML-Multiverse achieves this by explicitly retaining and interrogating the full distribution of pipelines generated during automated search. This enables systematic examination of where and why modelling conclusions diverge, rather than obscuring disagreement through aggregation.

We use the autoML-Multiverse to quantify analytical variability and use AD as an example of its biomedical application. We systematically investigate the stability and robustness of ML models across a range of classification tasks. Our primary aim is to assess which results persist across analytical configurations, rather than maximising accuracy, and thereby provide an empirical basis for instability-aware and robustness-focused evaluation in ML research. By explicitly characterising how modelling choices influence results, we seek to promote uncertainty-aware evaluation practices that support more robust, transparent, and clinically meaningful ML studies in AD and related diseases.

## 2. Materials and Methods

Using data from two independent AD cohorts, we applied the AutoML-Multiverse framework separately within each cohort across 20 prediction tasks, spanning diagnostic, disease-staging, and progression settings. Analyses were performed using imaging, clinical/cognitive, fluid biomarker, and multimodal feature configurations, allowing us to assess how conclusions vary across modelling choices, data sampling, data modalities, and cohorts.

### 2.1 Datasets and participants

Data used in the preparation of this article were obtained from the Alzheimer’s Disease Neuroimaging Initiative (ADNI) database (adni.loni.usc.edu). The ADNI was launched in 2003 as a public-private partnership, led by Principal Investigator Michael W. Weiner, MD. The primary goal of ADNI has been to test whether serial magnetic resonance imaging (MRI), positron emission tomography (PET), other biological markers, and clinical and neuropsychological assessment can be combined to measure the progression of mild cognitive impairment (MCI) and early Alzheimer’s disease (AD).

ADNI includes cognitively normal older adults, individuals with MCI, and patients with AD, assessed using standardised diagnostic criteria and harmonised acquisition and assessment protocols across sites. For the present analyses, ADNI data were accessed via the TADPOLE (The Alzheimer’s Disease Prediction Of Longitudinal Evolution) challenge dataset, which provides a curated subset of ADNI variables commonly used for disease classification and progression modelling. Diagnostic labels and task definitions followed TADPOLE conventions(25).

For each prediction task, we defined modality-specific feature sets (imaging, clinical/cognitive, and multimodal combinations) based on the available variables in the dataset. For ADNI, a set of participants was selected for each task (e.g., AD vs. CN) and those same participants were used across the three different experiments (imaging only, clinical/cognitive only, multi-modal). Detailed sample sizes and demographic characteristics for all ADNI tasks are reported in Table 1.

**Table 1.** Sample characteristics for ADNI tasks. Abbreviations: CN = cognitively normal; MCI = mild cognitive impairment; AD = dementia due to Alzheimer’s disease; sMCI = stable MCI; pMCI = progressive MCI; MMSE = Mini-Mental State Examination.

| Task | Modalities | N (per class) | Age (years), mean ± SD | Sex, n (%) | MMSE, median (IQR) |
| --- | --- | --- | --- | --- | --- |
| **CN vs. AD** | Imaging, Cognitive, Multimodal | 760 / 303 | 72.27 ± 7.4 (CN)<br>74.19 ± 8.0 (AD) | 366 (42.2%) F<br>135 (44.6%) F | 29.0 [1.0] (CN)<br>23.0 [4.0] (AD) |
| **CN vs. MCI vs. AD** | Imaging, Cognitive, Multimodal | 760 / 867 / 303 | 72.27 ± 7.4 (CN)<br>72.19 ± 6.5 (MCI)<br>74.19 ± 8.0 (AD) | 426 (56.1%) F<br>366 (42.2%) F<br>135 (44.6%) F | 29.0 [1.0] (CN)<br>27.5 [3.0] (MCI)<br>23.0 [4.0] (AD) |
| **sMCI vs. pMCI** | Imaging, Cognitive, Multimodal | 190 / 88 | 70.43 ± 7.0 (sMCI)<br>72.46 ± 6.8 (pMCI) | 75 (39.5%) F<br>36 (40.9%) F | 28.5 [1.0](sMCI)<br>28.2 [2.0](pMCI) |

Data for independent replication and cross-cohort evaluation were obtained from the National Alzheimer’s Coordinating Center (NACC) database, which aggregates uniformly collected clinical, cognitive, neuroimaging, and biomarker data from Alzheimer’s Disease Centers across the United States. Feature modalities for NACC analyses included imaging, clinical/cognitive variables, fluid biomarkers, and multimodal combinations, defined according to the task-specific configurations described below. The NACC cohort includes cognitively normal individuals, participants with MCI, and individuals with dementia due to suspected or probable AD, evaluated using standardised diagnostic criteria and longitudinal follow-up procedures.

Because each modality configuration (imaging, clinical/cognitive, fluid biomarkers, multimodal) requires a distinct set of variables, complete-case inclusion criteria were applied separately within each configuration. Consequently, sample sizes and analysis cohorts differ across modalities within the same prediction task. These modality- and task-specific sample sizes for NACC are reported in Table 2.

**Table 2.** Sample characteristics for NACC tasks. Abbreviations: CN = cognitively normal; MCI = mild cognitive impairment; AD = dementia due to Alzheimer’s disease; sMCI = stable MCI; pMCI = progressive MCI; CSF = cerebrospinal fluid; MoCA = Montreal Cognitive Assessment.

| Task | Modality | N (per class) | Age (years), mean ± SD | Sex, n (%) | MoCA, median (IQR) |
| --- | --- | --- | --- | --- | --- |
| **CN vs. MCI** | Imaging, Cognitive, Multimodal | 341 / 374 | 72.47 ± 7.2 (CN)<br>71.88 ± 7.9 (MCI) | 211 (61.9%) F<br>192 (51.3%) F | 27.3 [4.0]<br>23.3 [5.0] |
| **CN vs. MCI** | CSF | 850 / 207 | 71.04 ± 8.3 (CN)<br>72.09 ± 8.3 (MCI) | 346 (42.9%) F<br>117 (62.9%) F | N/A |
| **MCI vs. AD** | Imaging, Cognitive, Multimodal | 374 / 238 | 71.88 ± 7.9 (MCI)<br>70.66 ± 9.3 (AD) | 192 (51.3%) F<br>140 (58.8%) F | 23.3 [5.0]<br>19.0 [7.7] |
| **MCI vs. AD** | CSF | 207 / 564 | 72.09 ± 8.3 (MCI)<br>72.61 ± 9.0 (AD) | 117 (62.9%) F<br>288 (54.0%) F | N/A |
| **sMCI vs. pMCI** | Imaging | 106 / 119 | 72.69 ± 7.8 (sMCI)<br>74.29 ± 8.8(pMCI) | 46 (43.4%) F<br>57 (47.9%) F | N/A |
| **sMCI vs. pMCI** | Cognitive, Multimodal | 51 / 41 | 71.38 ± 5.9 (sMCI)<br>74.13 ± 6.9(pMCI) | 19 (37.3%) F<br>20 (48.8%) F | 23.1 [3.0]<br>21.7 [4.0] |

For progression prediction tasks, stable and progressive MCI were defined based on diagnosis status at three-year follow-up, applied consistently across both ADNI and NACC.

All data used in this study were obtained from the ADNI and NACC databases. Data were fully de-identified, and ethical approval and informed consent were obtained by the original study investigators at each participating site. The present analyses involved secondary use of these data and were conducted in accordance with the data use agreements and governance frameworks of ADNI and NACC.

### 2.2 Feature Modalities

#### 2.2.1 Structural MRI features

Structural imaging features were derived from T1-weighted MRI scans for both ADNI and NACC. Regional structural measures were extracted using FreeSurfer (26) (automated cortical and subcortical segmentation) and comprised regional volumetric features treated as continuous variables. Volumes were adjusted for intracranial volume (ICV) using linear regression–based residual adjustment within each dataset. All continuous features were standard scaled (zero mean, unit variance), with scaling parameters estimated on the training data within each resampling split and applied to the corresponding test data within the same cohort. For each task and modality configuration, all modelling approaches were trained and evaluated using identical feature sets and participant subsets, ensuring fair within-task comparisons. The complete lists of regional structural MRI features used for ADNI and NACC analyses are provided in the Supplementary Methods (Section S1.1; Tables S1.1 and S1.2).

#### 2.2.2 Clinical and cognitive measures

Clinical/cognitive features comprised age, sex, and a global cognitive screening measure: Mini-Mental State Examination (MMSE) for ADNI and the Montreal Cognitive Assessment (MoCA) for NACC. Age and cognitive scores were treated as continuous variables, and sex as a binary categorical variable.

#### 2.2.3 Fluid biomarkers (CSF)

Fluid biomarker features were considered as a separate modality in the NACC analyses and comprised CSF measures of amyloid-β (CSFABETA), phosphorylated tau (CSFPTAU), and total tau (CSFTTAU) (continuous variables). Fluid biomarker features were incorporated in the NACC analyses as a predefined modality-specific configuration to examine their contribution within that cohort. While CSF measures are also available in ADNI, the present study was not designed as a comprehensive cross-cohort biomarker comparison. Instead, modality configurations were defined within each cohort to illustrate instability-aware evaluation under controlled experimental settings. Accordingly, fluid biomarkers were evaluated within NACC in a cohort-specific context while maintaining internally consistent task–modality comparisons.

#### 2.2.4 Multimodal feature configurations

Multimodal feature configurations were defined a priori by combining structural MRI features with clinical/cognitive measures within a single modelling framework. In this study, *multimodal* therefore refers specifically to imaging + clinical/cognitive combined, rather than fusion of all available data types. Multimodal analyses were conducted for both ADNI and NACC. Fluid biomarker features were treated as a separate modality and were not included in multimodal configurations, allowing modality-specific effects to be examined without confounding due to cohort-specific biomarker inclusion.

### 2.3 AutoML-Multiverse Framework

The AutoML-Multiverse is a model-agnostic framework which defines a structured prediction space composed of a large set of candidate ML pipelines (∼ 20,000 preprocessing and modelling combinations). Pipelines are represented in a low-dimensional latent configuration space derived from prediction similarity, which is efficiently explored using Bayesian optimisation. The framework can also be used to construct data-driven ensembles by combining complementary pipelines. An overview of the framework is shown in Figure 1, which summarises its four sequential phases - latent pipeline representation, guided pipeline exploration, ensemble construction, and evaluation - and illustrates the latent configuration space used for pipeline exploration.

**Figure 1.**
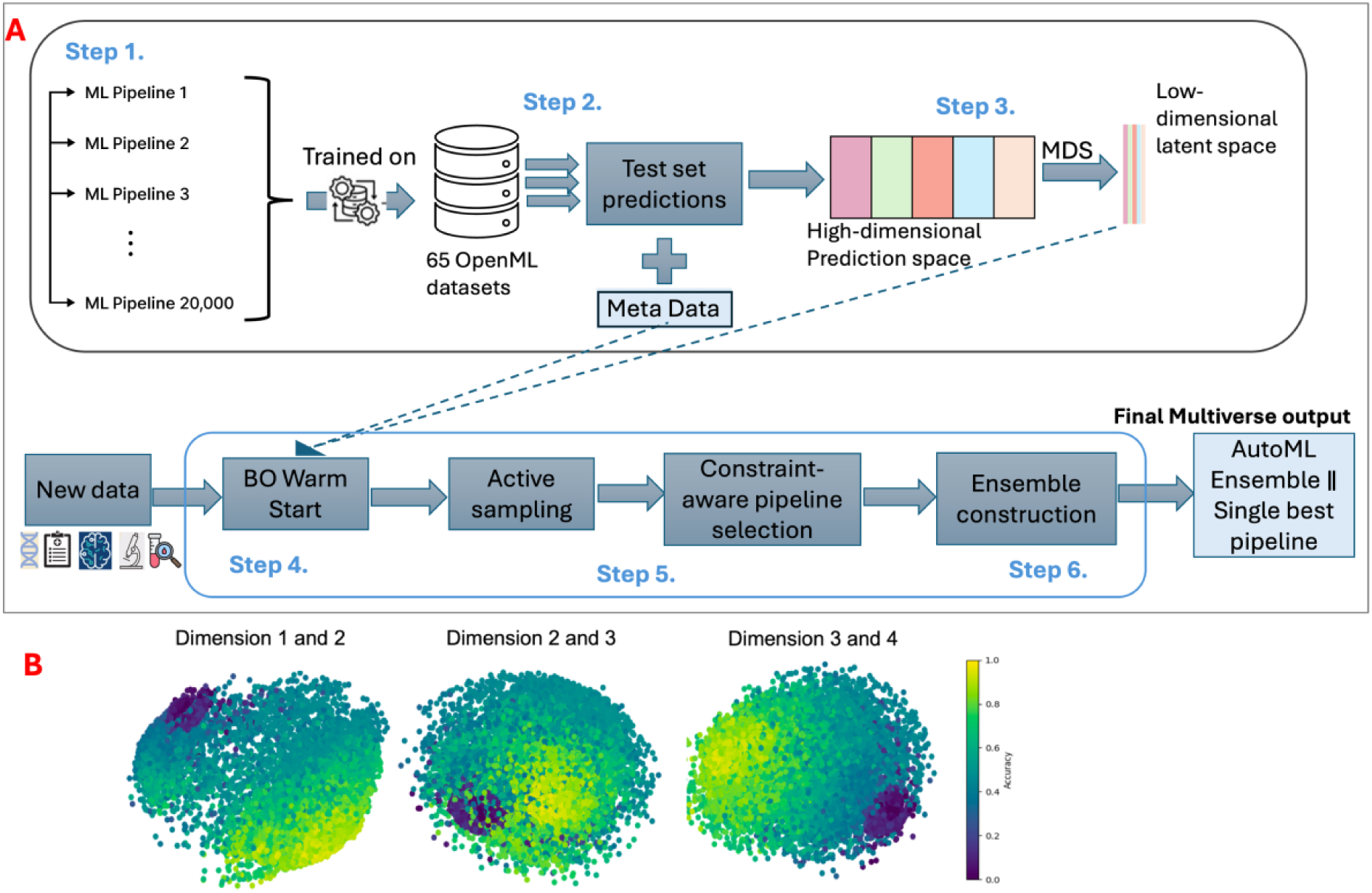
AutoML-Multiverse framework overview. **(A)** Schematic overview of the AutoML-Multiverse framework, illustrating the definition of a latent ML pipeline space, Bayesian optimisation with warm-start initialisation, and the construction of prediction-similarity-based ensembles from a large set of candidate modelling pipelines. **(B)** Conceptual visualisation of the low-dimensional latent configuration space, in which individual pipelines are represented as points positioned according to prediction similarity; proximity in the latent space reflects similarity in predictive behaviour across reference datasets.

AutoML searches over complete end-to-end pipelines, jointly optimising (i) missing-data imputation, (ii) feature preprocessing/encoding, (iii) the predictive model, and (iv) associated hyperparameters. In our configuration, each pipeline is drawn from a predefined component library comprising 5 imputation methods, 12 preprocessing operators (numerical/categorical), and 18 classifiers, with ∼82 tunable hyperparameters across components (Supplementary Material, Table X).

Full technical details of the latent space construction, warm-start strategy, Bayesian optimisation procedure, and ensemble generation algorithm are provided in the Supplementary Methods, section S1.2.

#### 2.3.1 Latent configuration space of pipelines

AutoML-Multiverse defines a high-dimensional space of analysis pipelines spanning data preprocessing, model specification, and hyperparameter configurations. In this space, proximity between pipelines reflects similarity in individual-level predictions rather than similarity in implementation details or overall accuracy. A conceptual illustration of this latent space and the relationship between pipeline proximity and predictive behaviour is provided in Figure 1B.

The latent configuration space is constructed by embedding pipelines based on their prediction similarity across a collection of reference datasets drawn from OpenML(27). Pipelines that generate similar predictions across multiple datasets are positioned close together, whereas pipelines exhibiting divergent predictive behaviour are separated in the latent space.

This representation provides a basis for navigating the modelling decision space that emphasises diversity among pipelines and is not constrained by predefined algorithm taxonomies. Consequently, pipelines from mathematically distinct model families (e.g., random forests and support vector machines) may occupy nearby regions of the latent space if they exhibit similar predictive behaviour.

#### 2.3.2 Guided pipeline exploration via Bayesian optimisation

Pipeline exploration in the latent space uses Bayesian optimisation (BO) with Gaussian process regression (28). The search is initialised using a warm-start strategy, in which candidate pipelines are ranked based on previously observed predictive performance and computational cost, and a subset of high-ranking pipelines is selected as initial seeds. BO then iteratively proposes new pipelines by balancing exploration of under-sampled regions of the configuration space with exploitation of regions associated with strong predictive performance.

To ensure computational efficiency, candidate pipeline proposals are constrained using metadata describing the computational cost and performance characteristics of nearby pipelines in the latent space. At each iteration, the optimisation process prioritises pipelines that are both proximal in the latent space to promising configurations and expected to be computationally tractable. This resource-aware strategy enables efficient traversal of a large and diverse modelling space while remaining feasible for repeated resampling-based evaluation.

#### 2.3.3 Ensemble construction and model retention

AutoML-Multiverse can construct data-driven ensembles by iteratively combining complementary pipelines identified during optimisation. Candidate pipelines are ranked according to validation performance, and ensemble construction proceeds in a greedy manner. At each step, a candidate pipeline is retained if it (i) improves validation performance by at least 1**%** relative to the current ensemble and (ii) exhibits sufficient prediction diversity with respect to the existing ensemble, quantified using Hamming distance.

Ensemble growth is terminated if no candidate pipeline yields further improvement after 20 consecutive iterations. When more than one pipeline is retained, the final predictive model is implemented as a stacked ensemble(29); otherwise, the single retained pipeline is used. This procedure ensures that retained models contribute complementary predictive information rather than redundancy.

### 2.4 Evaluation strategy

For each task–modality configuration, the framework was applied using identical feature sets and participant subsets across all modelling approaches, enabling direct comparison with individual ML models and stacked ensemble baselines. Model evaluation was designed to ensure fair and comparable assessment of classification performance across modelling strategies, data modalities, and cohorts.

All analyses were conducted using repeated, stratified train-test resampling, with a focus on evaluating model stability and generalisability rather than optimising performance under a single data split. For each prediction task and modality configuration, performance was assessed across 100 independent runs, defined as repeated random train-test resampling without fixed random seeds. In each run, data were split into 75% training and 25% testing sets, stratified by class label to preserve class balance. The 100 train-test splits were preserved across all modelling approaches within a given task-modality configuration, ensuring that performance differences reflected modelling strategy rather than variation in data partitioning.

Classification performance was quantified using balanced accuracy, computed on unseen test sets and summarised as the mean and standard deviation across the 100 runs. No model selection or optimisation was performed on test data.

For the AutoML-Multiverse, we additionally recorded, for each task and modality, the top five pipelines contributing to ensemble models across resampling runs to examine pipeline diversity and recurrence. These summaries accompany performance results to support interpretation of robustness and analytical flexibility.

#### 2.4.1 Benchmark modelling approaches

As benchmarks, a common set of modelling approaches was evaluated across all tasks and modalities. These included nine baseline ML models, a stacked ensemble model, and the proposed AutoML-Multiverse framework. The stacked ensemble followed a two-tier architecture, using the same nine individual models as base learners and a trainable Gaussian Naïve Bayes classifier(30) as the meta-learner. In this stacked ensemble, base-model predictions were used as inputs to the meta-learner, which was trained to combine them into a final prediction. All models were trained and evaluated using identical feature sets and participant subsets within each task-modality configuration. The baseline models evaluated are listed in the Supplementary Methods, section S1.3, Table S1.3 and details about stacked ensemble model are provided in section S1.3.1.

### 2.5 Statistical analysis

This study adopted a descriptive, resampling-based evaluation strategy rather than formal hypothesis testing. The primary objective was not to establish statistically significant performance differences between individual models, but to characterise the variability, stability, and robustness of ML-derived conclusions across equally valid analytical configurations.

Predictive performance was summarised using balanced accuracy, reported as the mean and standard deviation across 100 repeated train–test resampling runs for each task, modality, and modelling approach. Balanced accuracy was chosen to account for class imbalance, and variability across resampling runs was treated as an informative outcome in its own right, reflecting sensitivity to data partitioning and analytical choices.

Model comparisons were therefore conducted descriptively, with emphasis placed on the overlap, dispersion, and persistence of performance patterns across models, modalities, and cohorts, rather than on identifying a single “winning” model. Formal hypothesis testing and p-value–based comparisons were not performed, as the study was not designed to test predefined statistical hypotheses nor to claim statistically significant superiority of one modelling approach over another. Instead, the analysis provides an empirical characterisation of modelling instability and robustness, with the aim of supporting more transparent, reliable, and interpretable ML practice in Alzheimer’s disease research.

### 2.6 Software and implementation

All analyses were implemented in Python using Jupyter notebooks. Model development and evaluation were conducted using standard ML libraries, including *scikit-learn* (31) for classical models and ensemble methods, and *XGBoost* for gradient-boosted tree models(32).

All modelling pipelines, resampling procedures, and evaluation steps were implemented consistently across tasks, modalities, and cohorts.

## 3. Results

Across 20 diagnostic, prognostic, and disease-staging prediction settings in ADNI and NACC, AutoML-Multiverse achieved the highest grand-average balanced accuracy across tasks and modalities (0.723 ± 0.038), marginally above the stacked ensemble (0.717 ± 0.041) and all individual baselines. However, many task-level comparisons exhibited overlapping performance distributions across modelling approaches (e.g., ADNI AD vs. CN imaging). Detailed results are presented below, structured to summarise overall performance, task-level variability, modality effects, and AutoML-Multiverse pipeline behaviour.

### 3.1 AutoML performance across classification tasks

AutoML-Multiverse consistently achieved high accuracy relative to individual baselines and stacked ensembles (Figure 2). When performance was aggregated across all 20 tasks, AutoML-Multiverse yielded the highest overall mean balanced accuracy, with variability comparable to or lower than that observed for alternative modelling approaches.

**Figure 2.**
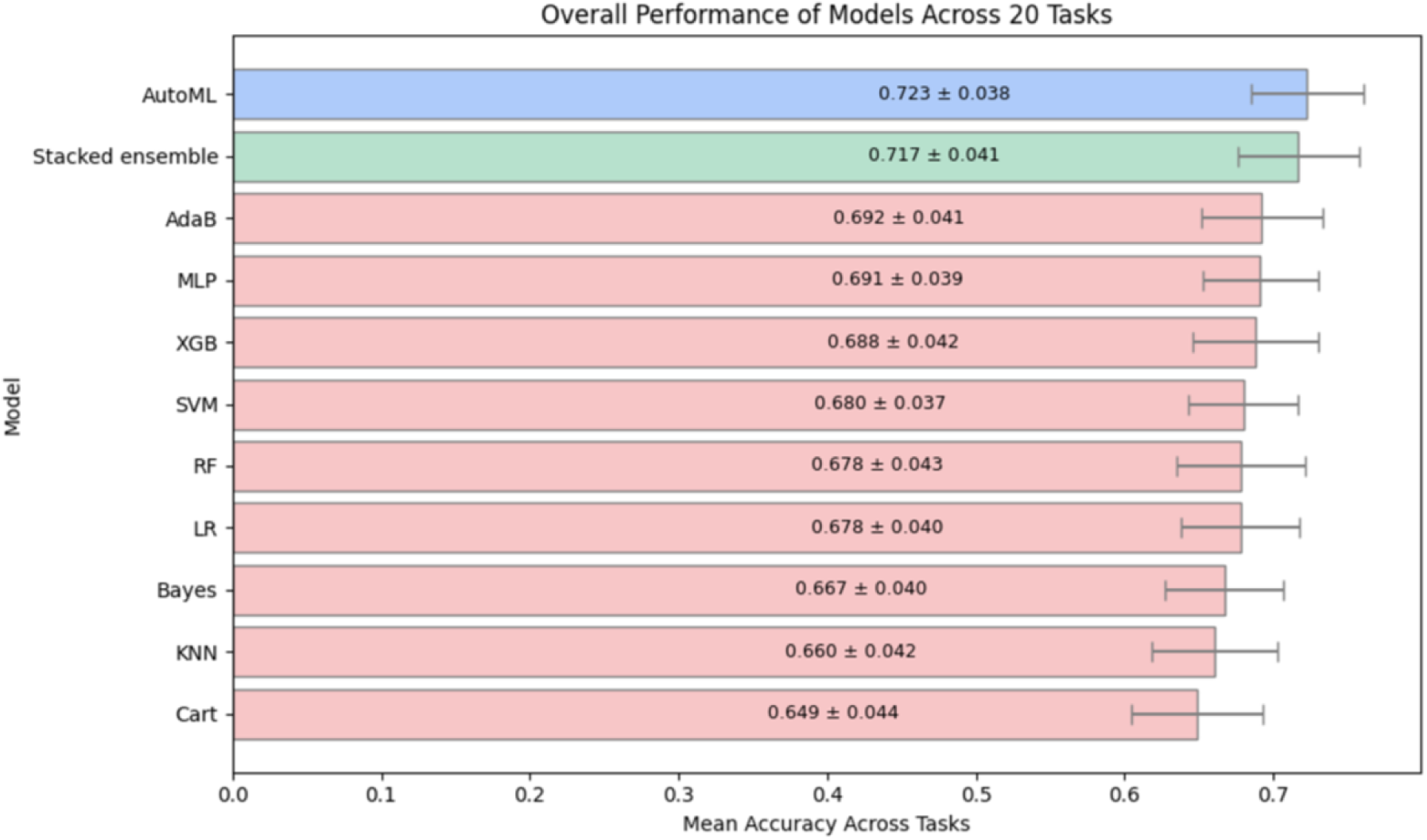
Grand-average predictive performance across tasks. Mean balanced accuracy (± standard deviation) aggregated across all 20 prediction settings spanning diagnostic, prognostic, and disease-staging tasks, data modalities, and cohorts. Performance is reported for nine baseline ML models, a stacked ensemble of baseline models, and the AutoML-Multiverse framework, based on 100 independent resampling runs with evaluation on unseen test sets. The plot summarises overall performance reliability across heterogeneous clinical scenarios rather than task-specific optimality.

At the level of individual tasks, AutoML-Multiverse was the top-performing approach in 11 of 20 prediction tasks. In the remaining tasks, it ranked second in 2 tasks and third in 2 tasks. Because AutoML-Multiverse explores the pipeline space using guided search rather than exhaustive evaluation, the optimal pipeline for a given task may not always be selected within the available search budget; consequently, individual baseline models may occasionally achieve marginally higher mean performance in specific task–modality settings. In most task–modality configurations, differences in mean balanced accuracy between AutoML-Multiverse and the top-ranked alternative were small and within one standard deviation of the performance distribution estimated across 100 resampling runs, indicating substantial overlap between competing approaches.

Across the 20 prediction tasks, the identity of the top-performing model varied by task and cohort. AutoML-Multiverse achieved the highest mean balanced accuracy in 11 tasks, while the stacked ensemble was top-performing in 4 tasks (including one task in which it was tied with SVM). Individual baseline models achieved highest overall performance in only a small number of settings: MLP in 2 tasks, XGBoost in 2 tasks, and logistic regression in 1 task. These counts refer to overall highest mean balanced accuracy across all modelling approaches within each task. When autoML-Multiverse models did not show top performance, their performance was generally numerically close to the top model. Full task-level results, including mean balanced accuracy and variability across resampling runs, are provided in Supplementary Material section S2.1 including Table S2.1.

#### 3.1.1. Performance variability across resampling runs

Across prediction tasks and modelling approaches, variability across repeated train-test resampling runs was substantial (Table 3 and Figure 2), reflecting sensitivity of performance estimates to data partitioning. When aggregated across tasks, the standard deviation of balanced accuracy ranged from 0.037-0.044. Notably, this magnitude of resampling variability is larger than many of the mean performance differences used to rank models within tasks. In other words, the partitioning of the data into training and test often had a bigger impact on performance than choice of model.

**Table 3.**
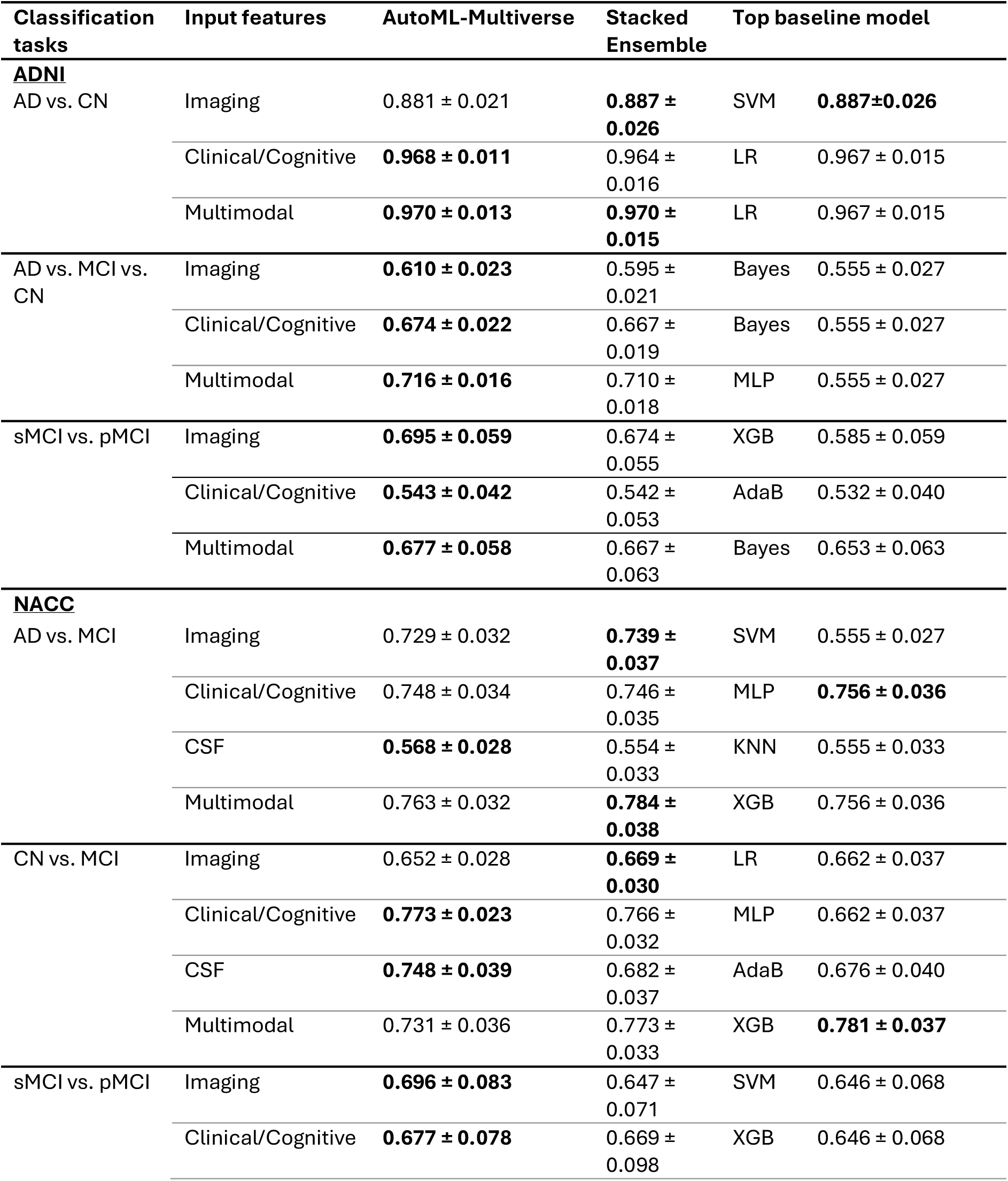
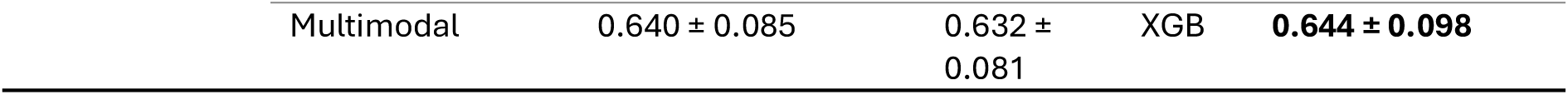
Summary of task-level predictive performance across 20 Alzheimer’s disease prediction settings in ADNI and NACC. Mean balanced accuracy (± standard deviation across 100 resampling runs) on unseen test sets is shown for AutoML-Multiverse, the stacked ensemble, and the most accurate individual baseline model for each task. Numerically highest accuracy scores per tasks are in bold. Full results for all evaluated baseline models are provided in Supplementary Material, Section S2.1, Table S2.1. AD = Alzheimer’s Disease, CN = Cognitively Normal, sMCI = stable Mild Cognitive Impairment, pMCI = progressive Mild Cognitive Impairment.

### 3.2 Instability across pipelines in AD classification and in MCI progression classification tasks

To provide more detailed illustration of task-level behaviour, we focus on two representative settings: AD classification using ADNI data and MCI progression classification using NACC. Figure 3 presents results for the ADNI AD versus cognitively normal (CN) classification task using multimodal features. This task represents a setting with highly separable groups and near-ceiling performance. Across modelling approaches, mean balanced accuracy was consistently high (0.946-0.973), with standard deviations ranging from 0.013 to 0.019 across resampling runs. The numerically highest mean performance was observed for logistic regression (0.973 ± 0.014), closely followed by MLP and SVM (both 0.971), while AutoML-Multiverse and the stacked ensemble achieved 0.970. Differences between top-ranked methods and alternative approaches were small relative to the observed resampling variability, and performance distributions exhibited substantial overlap. Consequently, rank ordering in this task was sensitive to modelling configuration, despite identical feature sets, train–test splits, and evaluation procedures.

**Figure 3.**
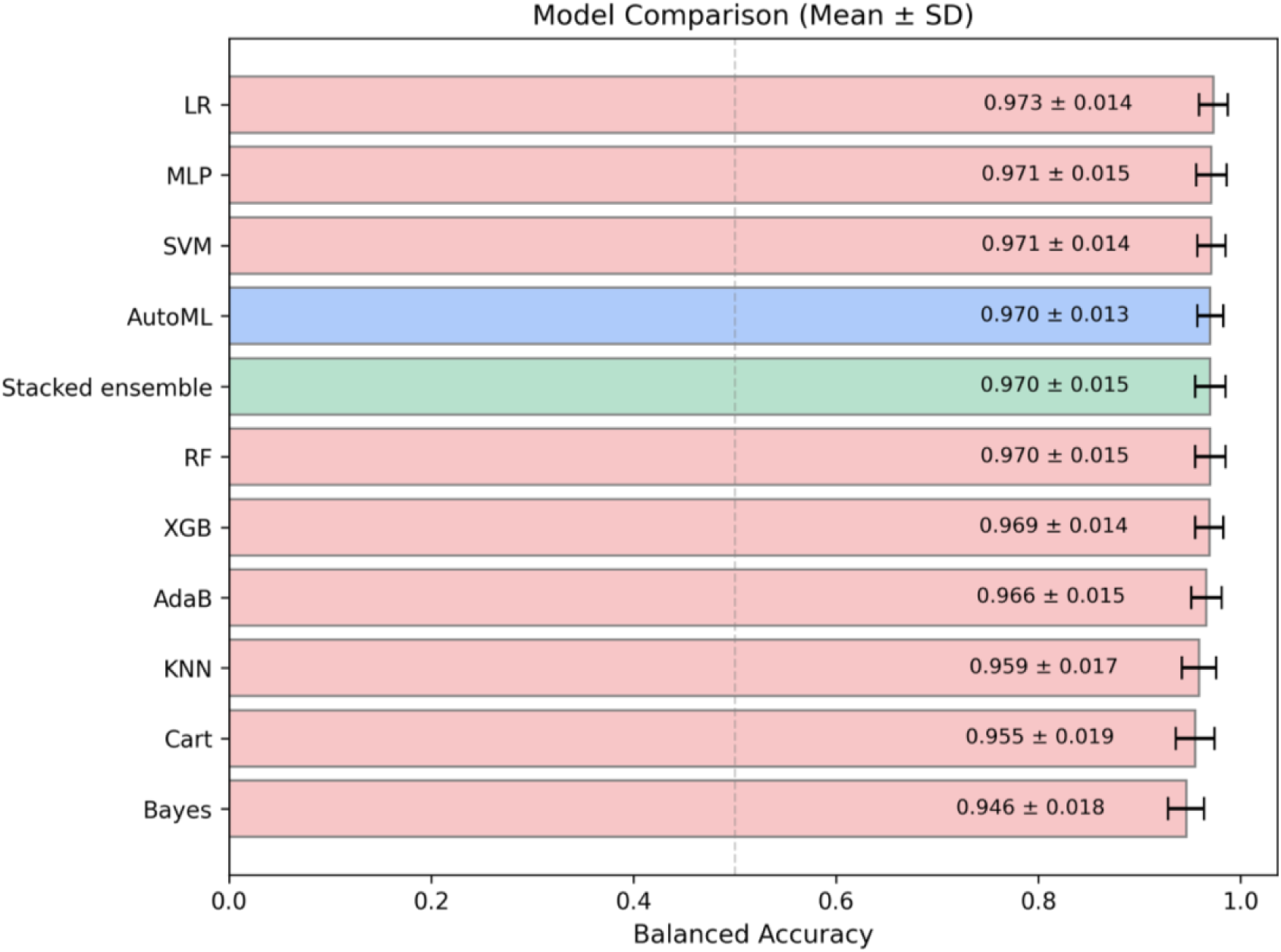
ADNI AD versus cognitively normal (CN) classification using multimodal features. Mean balanced accuracy (± standard deviation) across 100 independent resampling runs is shown for nine baseline ML models, a stacked ensemble of baseline models, and the AutoML-Multiverse framework, evaluated on unseen test sets.

Figure 4 presents the results for the NACC stable versus progressive mild cognitive impairment (sMCI versus pMCI) task using multimodal features. In this task the groups tend to be less separable, as they are clinically similar at time of baseline MCI assessment. Mean balanced accuracy again showed considerable overlap across all, with no single approach consistently separated from the others beyond the observed uncertainty. However, in contrast to AD versus CN, this task showed higher dispersion of performance estimates across resampling runs for all modelling approaches (standard deviations ranging from 0.081-0.107), showing greater sensitivity to data partitioning. Here, unlike the previous task, logistic regression (mean balanced accuracy = 0.596 ± 0.097) performed worse than the top baseline model (XGB mean balanced accuracy 0.644 ± 0.098), though given the high variability that difference might not be robust.

**Figure 4.**
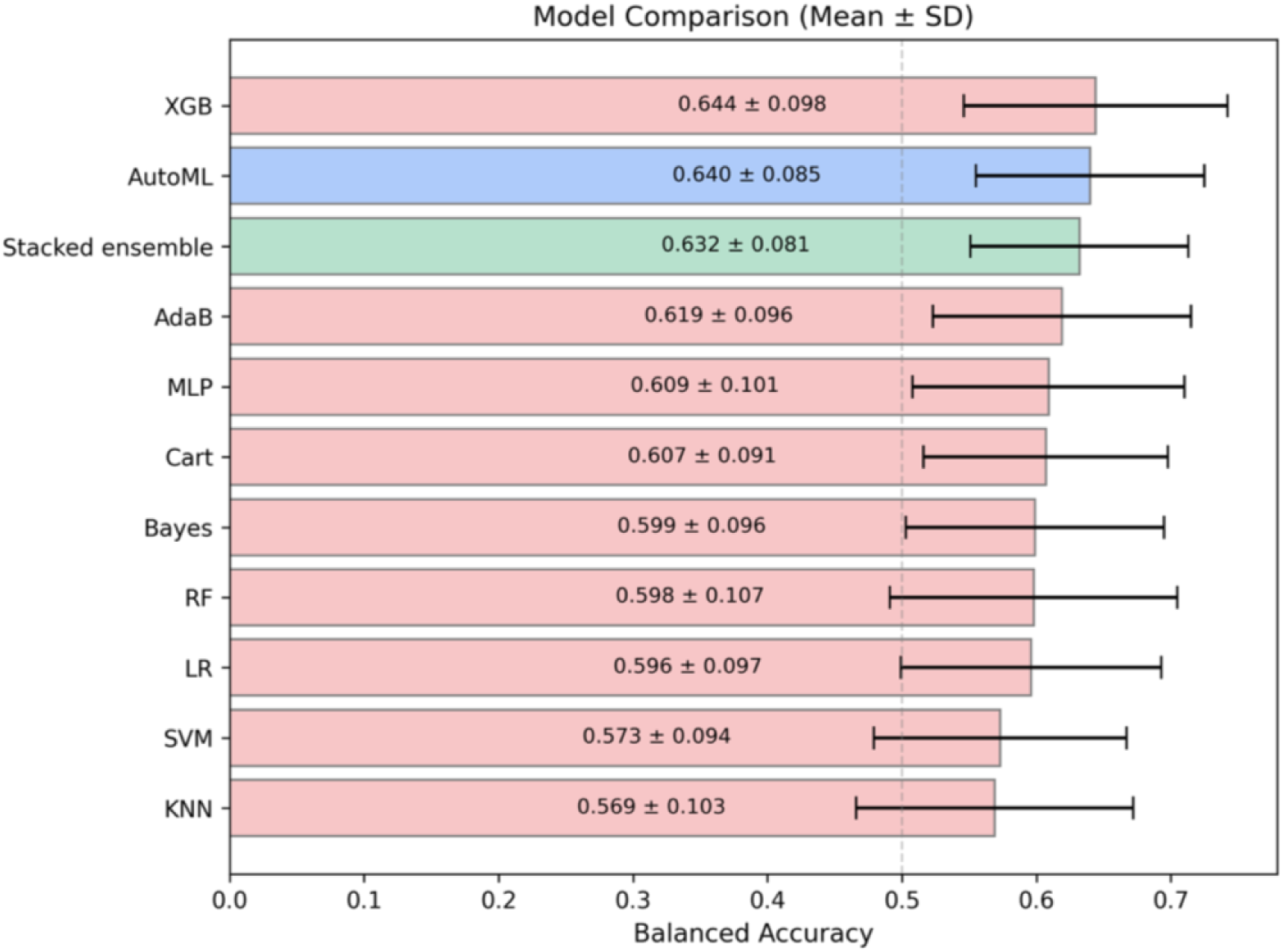
NACC stable versus progressive mild cognitive impairment (sMCI vs. pMCI) prediction using multimodal features. Mean balanced accuracy (± standard deviation) across 100 independent resampling runs is shown for baseline models, a stacked ensemble, and AutoML-Multiverse, evaluated on unseen test sets.

These examples demonstrate that it is hard to predict a priori which baseline model will give optimal accuracy. While the stacked ensemble performs comparatively well in both tasks, the more agnostic autoML-Multiverse approach can provide comparable results, without having to prespecify which algorithm to use.

### 3.3 Characterising autoML-Multiverse performance across tasks

Performance of the AutoML-Multiverse varied considerably across modalities and task definitions (Figure 5). For diagnostic tasks in which labels are defined based on cognitive performance (e.g., AD vs. CN), clinical/cognitive and multimodal configurations generally achieved higher balanced accuracy than imaging-only models. For example, in ADNI AD vs. CN classification, balanced accuracy ranged from 0.881 for imaging to 0.968-0.970 for clinical/cognitive and multimodal configurations. Across other cognitively defined diagnostic settings, performance typically ranged from approximately 0.67 to 0.77 for clinical/cognitive and multimodal inputs, compared with lower values for imaging-only configurations in some tasks.

**Figure 5.**
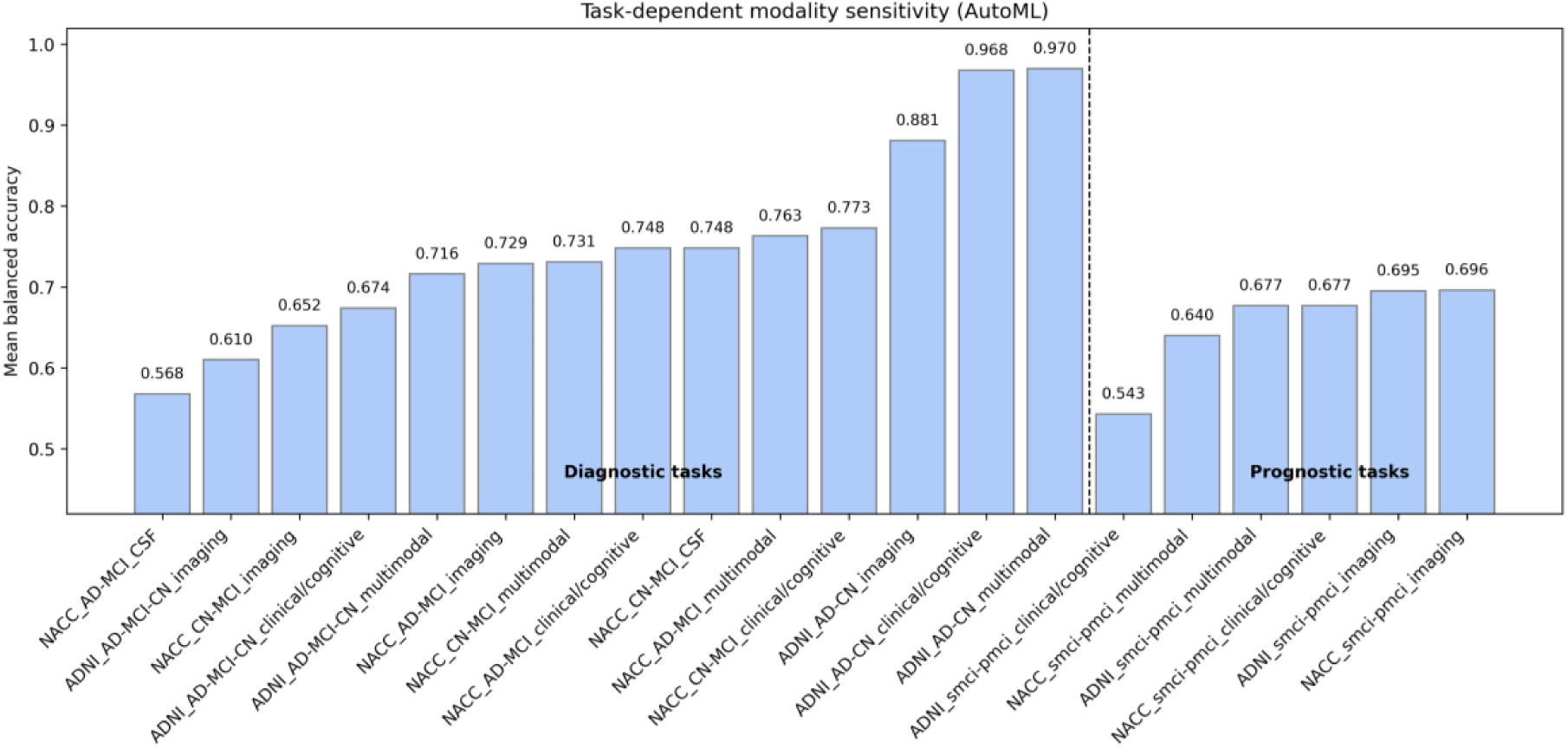
Shift in predictive utility across classification tasks. Mean balanced accuracy for AutoML-Multiverse is shown across imaging, clinical/cognitive, fluid biomarker, and multimodal configurations in ADNI and NACC. Diagnostic tasks exhibit higher separability, whereas prognostic tasks show reduced overall performance and greater task-dependent variability across modalities.

In contrast, for stable versus progressive MCI classification - where baseline cognitive information is more equivocal by definition - imaging-based models frequently performed comparably to or slightly better than clinical/cognitive configurations. In these prognostic tasks, balanced accuracy ranged from approximately 0.54 to 0.70, with imaging-based configurations (0.695-0.696) performing at or above clinical/cognitive models (0.543-0.677), and multimodal integration not uniformly improving performance.

### 3.4 Examining AutoML-Multiverse model selections across classification tasks

#### 3.4.1 Ensemble formation

To examine how AutoML-Multiverse selects models across classification tasks, we assessed how often ensembles were constructed (Figure 6). Ensembles were only constructed when it was automatically determined that they improved performance; in our results this occurred in a task and modality-dependent manner.

**Figure 6.**
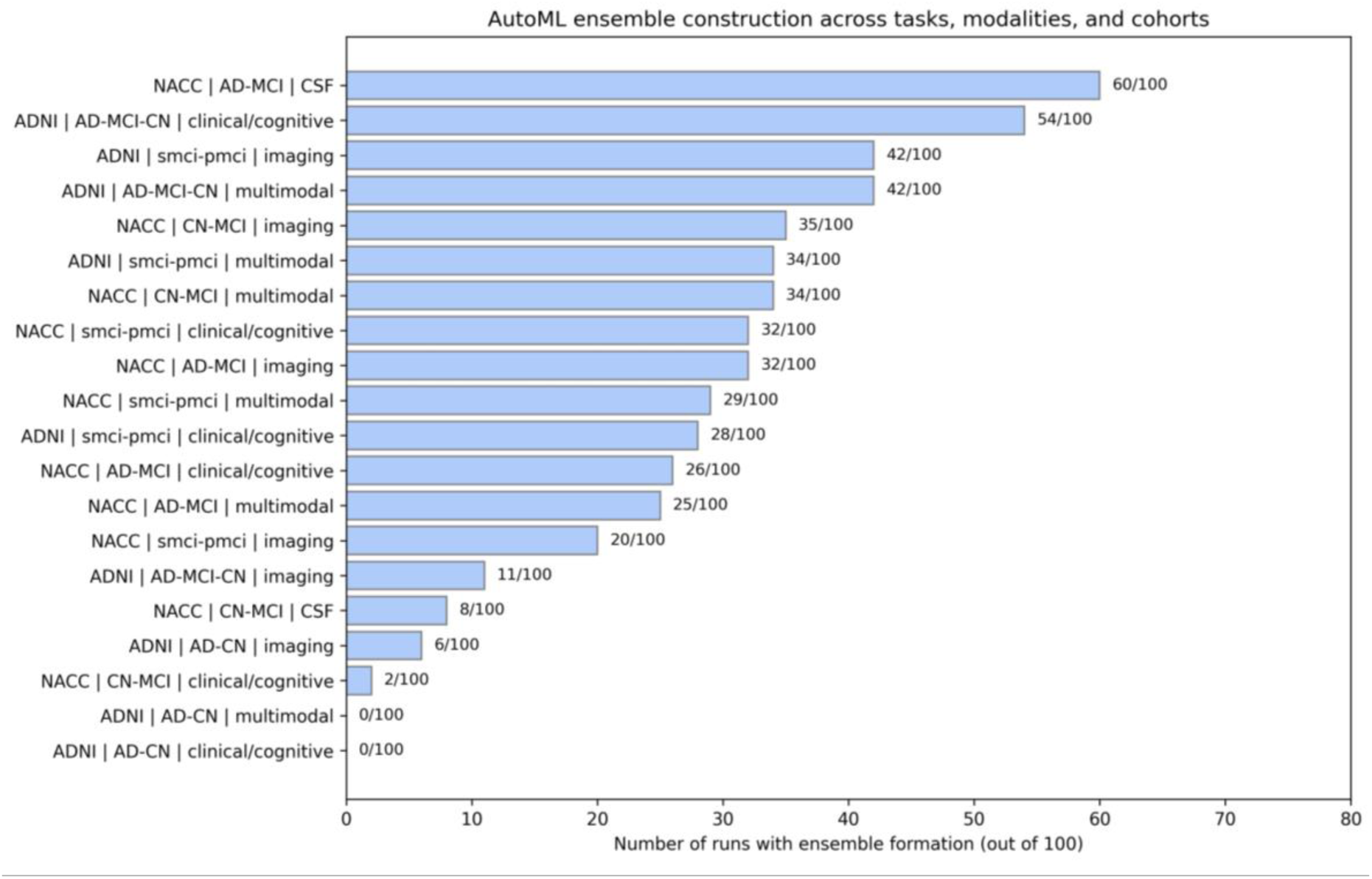
Cohort-, task-, and modality-dependent ensemble formation in AutoML-Multiverse. The number of resampling runs (out of 100) in which AutoML-Multiverse constructed an ensemble model is shown for each task-modality-cohort configuration. Ensemble formation was selective rather than ubiquitous, occurring more frequently in specific prognostic and multimodal settings. This pattern indicates that ensemble construction reflects underlying analytical plurality rather than being imposed uniformly across prediction tasks.

AutoML-Multiverse constructed ensemble models in some task-modality experiments but consistently selected single pipelines in others. Diagnostic classification tasks, particularly those typically more separable groups (e.g., AD vs. CN), tend to converge on a single pipeline. In contrast, several more challenging tasks (e.g., MCI progression or three label classification) more frequently resulted in ensemble construction.

Ensemble construction differed across modalities. Multimodal and imaging-based configurations more often resulted in ensembles compared to models using clinical/cognitive features only. Differences were also observed between ADNI and NACC, though this varied based on the experiment.

These findings reflect the adaptive design of the AutoML-Multiverse framework. Ensemble aggregation is not enforced by default; instead, ensembles are constructed only when multiple pipelines contribute complementary predictive information. As observed here, this resulted in ensemble solutions in some task-modality configurations but not others. A detailed summary of ensemble formation frequency and the most recurrent pipeline types across tasks and modalities is provided in Supplementary Table S2.2.

#### 3.4.2 Pipeline diversity persists across cohorts

To characterise pipeline diversity, we quantified the recurrence of individual pipelines among the top-performing models across resampling runs by assessing how often each pipeline appeared in the top five (ranked by mean balanced accuracy) across all 20 experiments (2000 AutoML-Multiverse runs). AutoML-Multiverse returned a diverse range of pipelines across both cohorts, with no single pipeline consistently dominating (Figure 7). The most frequently selected pipeline family comprised SVM-based configurations, appearing in 21.6% of runs (432 of 2000). These SVM-based pipelines reflect classifier components selected within the AutoML search space and are distinct from the standalone SVM baseline model evaluated separately. Overall, AutoML-Multiverse selected a heterogeneous mix of modelling strategies, including linear classifiers, kernel-based methods, and tree-based approaches. Diversity was also observed in upstream pipeline components, including imputation strategies and feature preprocessing operators, reflecting the broader AutoML search space over complete end-to-end pipelines (Supplementary Table S1.2.1).

**Figure 7.**
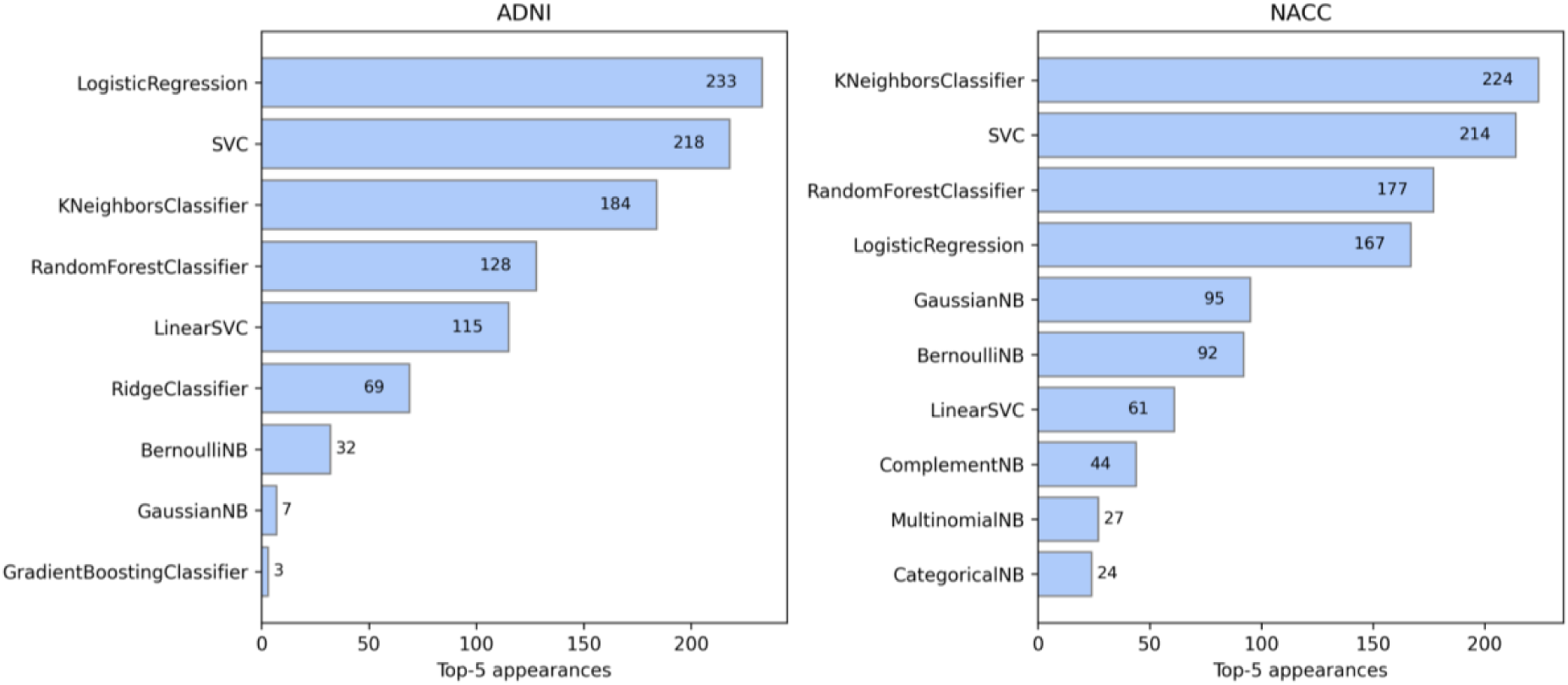
Pipeline selection frequency among top-performing AutoML-Multiverse models. Bars indicate the number of times individual pipelines appeared among the top five AutoML-Multiverse solutions across resampling runs, shown separately for ADNI (9 experiments, thus 900 runs) and NACC (11 experiments, thus 1100 runs).

These results indicate that pipeline optimality varies across cohorts and resampling runs and is not trivially predictable *a priori*.

## 4. Discussion

In this study, we examined the stability of inferences drawn from ML evaluation results in AD, using the AutoML-Multiverse framework across 20 diagnostic, prognostic, and disease-staging tasks in two independent cohorts. For example, we can draw conclusions about which modelling approach appears best, which modality is most informative, and how sensitive these findings are to analysis design and resampling. Our aim is not to recommend a universal “best” algorithm, but to propose a framework that provides more reliable inference for a given research question by reducing researcher-induced biases and making sensitivity to analysis choices more explicit.

Rather than identifying a single best-performing model, we evaluated how conclusions about performance, modality utility, and analytic choices (e.g., algorithm family, hyperparameter configuration, and ensemble strategy) shift under equally defensible configurations and repeated resampling. Across tasks, we observed substantial overlap in performance distributions, sensitivity to data partitioning, and task-dependent reversals in model rankings. Within this instability-aware framework, AutoML-Multiverse maintained consistently competitive performance while explicitly characterising analytic variability, supporting a move away from leaderboard-style interpretations toward robustness- and uncertainty-aware evaluation in AD ML, with flexibility to adapt to the specific domain context without requiring a priori commitment to particular modelling choices.

### 4.1 Instability in ML-based Alzheimer’s disease research

Variability in conclusions drawn from ML evaluation results in AD arises not simply from flawed modelling or idiosyncratic datasets, but from the interaction between complex biological signal, heterogeneous clinical populations, finite sample sizes, and expansive analytical decision spaces. Across diagnostic, prognostic, and staging tasks, we observed substantial overlap in performance distributions, frequent reversals in model rankings, and sensitivity to data resampling - patterns that persisted across modalities and cohorts even under standardised protocols and identical inputs. The substantial overlap in task-level performance distributions implies that “best model” conclusions are often contingent on the analytic configuration chosen, rather than reflecting a clearly dominant solution.

This observation aligns with broader evidence from neuroimaging and computational neuroscience demonstrating that when multiple, equally defensible analytic choices exist, conclusions can vary meaningfully (8)(10)(33)(34)(9). In the context of clinical ML for AD, the analytical decision space is particularly expansive, encompassing model architecture, hyper-parameterisation, preprocessing strategies, data modality selection, and cohort composition. Our results show that navigating this space inevitably yields a flexibility of plausible outcomes rather than a single, definitive answer, even when evaluation is performed rigorously and transparently.

Importantly, instability was observed not only across modelling approaches but also within the same approach under repeated resampling. Variability induced by stochastic train-test partitioning contributed to uncertainty in estimated performance, indicating that single-split evaluations substantially underrepresent the range of plausible outcomes. Conceptually, this distinction can be framed in terms of aleatoric and epistemic uncertainty. Aleatoric variability reflects irreducible heterogeneity in disease expression and measurement noise. In contrast, epistemic variability arises from limited data, sampling variability, analytic flexibility, and characteristics of the available signal and data distribution, and is, at least in principle, reducible through improved modelling and expanded data. Our findings suggest that a meaningful component of observed performance instability reflects epistemic uncertainty. AutoML-Multiverse does not eliminate such variability but makes it explicit by characterising the distribution of viable analytical solutions rather than privileging a single pipeline. Reporting this variability provides a more informative basis for inference than relying on marginal differences in point estimates.

### 4.2 Implications for modality integration in clinical ML for AD

The modality-related findings of this study should be interpreted cautiously and within the scope of the evaluated feature configurations. Our results challenge the assumption that multimodal fusion necessarily yields more reliable or clinically informative predictions (35)(36).

Combining heterogeneous data sources can reduce effective sample size due to missingness constraints, introduce redundancy and collinearity, and increase dimensionality relative to available observations - factors known to impair generalisation in high-dimensional clinical ML settings. In neuroimaging ML contexts where feature dimensionality approaches effective sample size, these trade-offs may offset potential gains from additional modalities, particularly when signal-to-noise ratios differ across feature types.

Our results emphasise that modality selection in AD ML studies should be driven by the specific clinical question and evaluated under uncertainty-aware frameworks. Generalised claims regarding the superiority of particular data sources(37)(38) risk oversimplifying a complex landscape in which diagnostic, prognostic, and staging objectives impose distinct informational demands.

### 4.3 Conceptual and methodological contributions of AutoML-Multiverse

ML approaches, including automated ML frameworks, have increasingly been adopted in AD research as a means of reducing manual analytic effort and improving predictive performance. However, most applications in clinical settings continue to frame automation primarily as a performance-optimisation tool, emphasising identification of a single best-performing pipeline or ensemble (39)(40)(19). In contrast, the principal contribution of AutoML-Multiverse lies not in automation alone, but in its ability to systematically expose and characterise analytical flexibility across complex modelling spaces.

Rather than collapsing thousands of plausible pipelines into a single optimal solution, AutoML-Multiverse retains and interrogates variability across modelling choices, data sampling, modalities, and cohorts. By representing automated pipeline search as a distribution of outcomes rather than a single performance estimate, our framework enables direct assessment of robustness and stability. In this sense, AutoML-Multiverse reframes automation as a mechanism for structured scientific inference under uncertainty.

The adaptive behaviour observed within AutoML-Multiverse - manifested through selective ensemble formation and persistent pipeline diversity - further illustrates this principle. Ensembles were constructed only when multiple pipelines exhibited complementary predictive behaviour, whereas single pipelines were selected where convergence occurred. This selective aggregation reflects the structure of the latent analytical landscape - in which distance corresponds to similarity in individual-level predictions rather than overall accuracy - rather than algorithmic indecision. This highlights that ensemble utility is contingent on the task and data context and is difficult to predict a priori, motivating an evaluation framework that can remain agnostic to whether ensembling will be beneficial.

From a methodological perspective, AutoML-Multiverse provides a practical template for instability-aware evaluation that is agnostic to specific model classes or data types. Although instantiated here using structured ML pipelines, the underlying principles - explicit exploration of analytical decision spaces, distributional reporting of performance, and adaptive aggregation - are broadly applicable to other modelling paradigms, including deep learning and multimodal neuroimaging analyses.

While this study did not focus on feature attribution, the persistence of high-performing solutions across diverse pipelines suggests an opportunity for consensus-based interpretability. Agreement in selected features or feature families across heterogeneous models may provide more robust insight than explanations derived from a single pipeline. Exploring feature-level stability across multiverse solutions represents an important direction for future work.

### 4.4 Methodological implications for future Alzheimer’s disease ML studies

The findings of this study have important implications for the design and evaluation of ML analyses in AD research.

First, evaluation practices must move beyond single train-test splits and isolated point estimates of performance. Across tasks and cohorts, resampling-induced variability materially influenced estimated performance, indicating that distributional reporting should be considered a minimum standard for robust evaluation. Reporting only peak or single-split accuracy risks overstating confidence and obscuring clinically relevant uncertainty.

Second, predictive objectives differ in their informational demands. Methodological choices - including modality configuration and model class - should therefore be evaluated relative to the specific clinical question rather than assumed to generalise across diagnostic, prognostic, and staging settings. Study design should be explicitly aligned with the target clinical decision context.

Third, comparative ML studies should prioritise robustness and transferability over nominal performance rankings. Cross-cohort evaluation, where feasible, provides a safeguard against cohort-specific overfitting and should be interpreted alongside within-cohort stability analyses to assess generalisability.

Broadly, these considerations argue for evaluation strategies that emphasise robustness, uncertainty characterisation, and reproducibility, alongside performance. Aligning methodological practice with these principles is essential for ensuring that ML-based evidence in AD research meets the standards required for meaningful clinical inference.

### 4.5 Limitations and future directions

Several limitations of this study should be acknowledged. First, although we evaluated a broad range of tasks across two large cohorts, the specific feature sets and modality configurations analysed here reflect the data availability and preprocessing choices adopted in this work rather than the full scope of variables contained within ADNI and NACC. Alternative feature representations or harmonisation strategies may yield different performance profiles and warrant further investigation. In addition, CSF biomarkers were incorporated only in NACC as part of a modality-specific evaluation. Although CSF data are available in ADNI, they were not included in the ADNI analyses in this study. This reflects a design choice to maintain controlled comparisons within each task-modality configuration. Future work may examine harmonised fluid biomarker analyses across cohorts.

Second, this study adopted a descriptive, resampling-based evaluation strategy rather than formal hypothesis testing. The objective was to characterise variability and robustness across analytical configurations rather than establish statistically significant superiority between models. Complementary statistical frameworks for summarising uncertainty across large model ensembles or multiverse analyses represent an important direction for future work.

Third, the analyses focused on structured ML pipelines and did not include end-to-end deep learning models. Although deep learning approaches are increasingly used in neuroimaging research (41)(42), their limited interpretability and substantial computational demands (43)(44) present additional challenges for instability-aware evaluation. Extending multiverse-style frameworks to encompass deep learning architectures, representation learning strategies, and hybrid models and emerging foundation-model approaches represents an important direction for future research.

Although exploring large analytical spaces incurs an upfront computational expense, this investment may be outweighed by the downstream risks of deploying unstable or misleading models in clinical research settings. From this perspective, instability-aware evaluation represents a trade-off between computation and evidentiary reliability rather than an impractical burden.

Finally, while AutoML-Multiverse was applied here as an operational framework to surface analytical flexibility, the broader principles of instability-aware evaluation are not limited to this specific implementation. Future work could extend these concepts to additional clinical domains, longitudinal modelling strategies, and expanded modality configurations to further clarify how uncertainty propagates across analytical choices.

### 4.6 Conclusion

This study provides empirical evidence that variability in ML evaluation results in Alzheimer’s disease research is pervasive, systematic, and informative. By explicitly characterising this variability and reframing evaluation around robustness rather than optimisation alone, the AutoML-Multiverse framework offers a principled foundation for more reliable and transparent machine-learning research in biomedical settings. Models intended for clinical use must therefore not only perform well on average, but also demonstrate stability within their intended clinical context (i.e., a given task definition and data environment) across reasonable modelling choices and data perturbations to support trustworthy individual-level predictions.

## CRediT authorship contribution statement

**Maitrei Kohli**: Writing – original draft, Conceptualization, Methodology, Formal analysis, Funding acquisition, Project administration, Data curation, Visualization. **Gonzalo Castro Leal**: Data curation, Writing – review. **Douglas Wyllie**: Writing – review. **Neil Oxtoby**: Writing – review. **Robert Leech**: Methodology, Conceptualization, Writing – review. **Philip Weston**: Writing – review. **James Cole**: Conceptualization, Methodology, Supervision, Project administration, Resources, Funding acquisition, Writing – review and editing. All authors read, reviewed and approved the final manuscript.

## Funding

**Maitrei Kohli** was supported by a UCL Computer Science Early Career Researcher Mini Fellowship.

## Acknowledgements

Data collection and sharing for this project was funded by the Alzheimer’s Disease Neuroimaging Initiative (ADNI) (National Institutes of Health Grant U01 AG024904) and DOD ADNI (Department of Defense award number W81XWH-12-2-0012). ADNI is funded by the National Institute on Aging, the National Institute of Biomedical Imaging and Bioengineering, and through generous contributions from the following: AbbVie, Alzheimer’s Association; Alzheimer’s Drug Discovery Foundation; Araclon Biotech; BioClinica, Inc.; Biogen; Bristol-Myers Squibb Company; CereSpir, Inc.; Cogstate; Eisai Inc.; Elan Pharmaceuticals, Inc.; Eli Lilly and Company; EuroImmun; F. Hoffmann-La Roche Ltd and its affiliated company Genentech, Inc.; Fujirebio; GE Healthcare; IXICO Ltd.; Janssen Alzheimer Immunotherapy Research & Development, LLC.; Johnson & Johnson Pharmaceutical Research & Development LLC.; Lumosity; Lundbeck; Merck & Co., Inc.; Meso Scale Diagnostics, LLC.; NeuroRx Research; Neurotrack Technologies; Novartis Pharmaceuticals Corporation; Pfizer Inc.; Piramal Imaging; Servier; Takeda Pharmaceutical Company; and Transition Therapeutics. The Canadian Institutes of Health Research is providing funds to support ADNI clinical sites in Canada. Private sector contributions are facilitated by the Foundation for the National Institutes of Health (www.fnih.org). The grantee organization is the Northern California Institute for Research and Education, and the study is coordinated by the Alzheimer’s Therapeutic Research Institute at the University of Southern California. ADNI data are disseminated by the Laboratory for Neuro Imaging at the University of Southern California.

## Declaration of competing interest

The authors declare that they have no competing interests.

## Data Availability Statement

The data that support the findings of this study were obtained from the National Alzheimer’s Coordinating Center (NACC) and the Alzheimer’s Disease Neuroimaging Initiative (ADNI). These data were used under license and are not publicly available. Access to the data may be requested directly from NACC (https://naccdata.org) and ADNI (https://adni.loni.usc.edu) subject to their respective data use agreements and approval processes.

## Notes

### Competing Interest Statement

The authors have declared no competing interest.

### Funding Statement

This study did not receive any external funding

### Author Declarations

Ethics approval for the Alzheimer's Disease Neuroimaging Initiative (ADNI) and the National Alzheimer's Coordinating Center (NACC) studies was obtained by the respective study investigators at participating institutions. All participants provided written informed consent. The present study used de-identified data obtained through approved data access procedures from the ADNI and NACC databases. ADNI- https://adni.loni.usc.edu and NACC - https://naccdata.org

