## Supplementary Materials File for "AutoML-Multiverse: An Instability-Aware Framework for Quantifying Analytic Variability in Alzheimer’s Disease Machine-Learning Studies"

#### S1. Materials and Methods

#### S1.1 Datasets & Participants – structural MRI features

Structural MRI features were derived from T1-weighted scans processed using FreeSurfer(1) for both ADNI(2) and NACC(3) cohorts. For each dataset, a predefined subset of regional volumetric measures was selected for analysis based on availability in the preprocessed datasets used in this study. These feature sets reflect the variables included in the present analyses rather than the full range of imaging measures available within each cohort.

All volumetric measures were corrected for intracranial volume (ICV) prior to modelling, and all continuous features were standard scaled to zero mean and unit variance. The selected imaging features for ADNI and NACC are listed in Tables S1.1 and S1.2, respectively.

| Feature name | Anatomical region |
|---|---|
| **Ventricles** | Ventricular system |
| **Hippocampus** | Hippocampus |
| **WholeBrain** | Whole brain volume |
| **Entorhinal** | Entorhinal cortex |
| **Fusiform** | Fusiform gyrus |
| **MidTemp** | Middle temporal gyrus |

**Table S1.1. Structural MRI features used in ADNI analyses**. All features represent regional volumetric measures derived from T1-weighted MRI and corrected for intracranial volume.

| Cortical regional volumes | Subcortical and ventricular volumes |
|---|---|
| **Feature name** | **Feature name** |
| bankssts_volume | Amygdala |
| caudalanteriorcingulate_volume | BrainSegVolNotVent |
| caudalmiddlefrontal_volume | Caudate |
| cuneus_volume | Cerebellum_Cortex |
| entorhinal_volume | Cerebellum_White_Matter |
| frontalpole_volume | Fourth_Ventricle |
| fusiform_volume | Hippocampus |
| inferiorparietal_volume | Inf_Lat_Vent |
| inferiortemporal_volume | Lateral_Ventricle |
| insula_volume | Pallidum |
| isthmuscingulate_volume | Putamen |
| lateraloccipital_volume | Thalamus |
| lateralorbitofrontal_volume | Third_Ventricle |
| lingual_volume | VentralDC |
| medialorbitofrontal_volume | |
| middletemporal_volume | |
| paracentral_volume | |
| parahippocampal_volume | |
| parsopercularis_volume | |
| parsorbitalis_volume | |
| parstriangularis_volume | |
| pericalcarine_volume | |
| postcentral_volume | |
| precentral_volume | |
| precuneus_volume | |
| posteriorcingulate_volume | |
| rostralanteriorcingulate_volume | |
| rostralmiddlefrontal_volume | |
| superiorfrontal_volume | |
| superiorparietal_volume | |
| superiortemporal_volume | |
| supramarginal_volume | |
| temporalpole_volume | |
| transversetemporal_volume | |

**Table S1.2. Structural MRI features used in NACC analyses**. All features represent regional volumetric measures derived from T1-weighted MRI and corrected for intracranial volume.

#### S1.2 AutoML-Multiverse framework: technical details

This section is intended to provide sufficient detail to enable conceptual understanding and methodological reproducibility, rather than to prescribe a specific software implementation. Here, we provide the full technical specification of the AutoML-Multiverse framework, expanding on the overview presented in the main Methods. AutoML-Multiverse is designed to efficiently explore a large space of candidate machine-learning pipelines while retaining information about model plurality and variability.

Figure S1.2 presents a schematic overview of the framework, illustrating the construction of the latent pipeline configuration space, the warm-started Bayesian optimisation strategy used to guide pipeline exploration, and the procedure for ensemble generation based on prediction similarity. The subsections below describe each component in detail, including latent space construction, optimisation dynamics, stopping criteria, and ensemble formation.

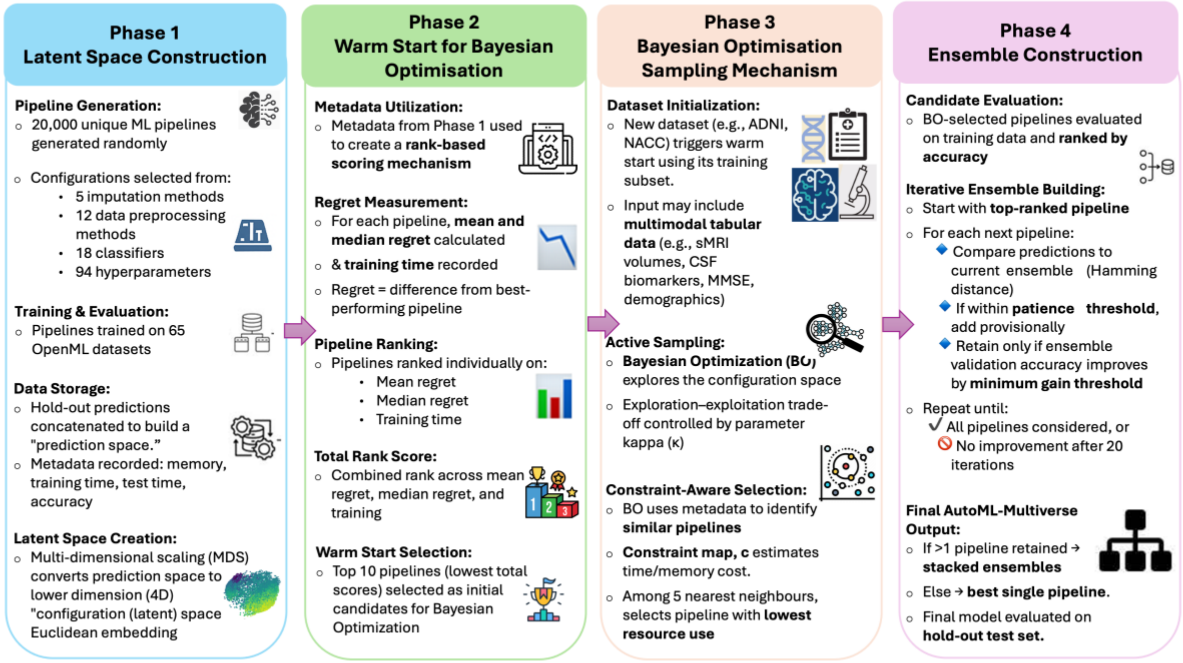


**Figure S1.2: Detailed schematic of the AutoML-Multiverse framework.** Overview of the four sequential phases of the AutoML-Multiverse framework: **(Phase 1)** construction of a latent configuration space from a large set of candidate machine-learning pipelines using prediction similarity; **(Phase 2)** warm-start initialisation based on pipeline metadata and regret-based ranking; **(Phase 3)** Bayesian optimisation–guided exploration of the latent space with resource-aware constraints; and **(Phase 4)** iterative ensemble construction based on prediction diversity and validation performance. Parameter values and implementation details are described in the following paragraphs.

### (Phase 1): Creating low dimensional latent space

**Objective:** To map diverse ML pipelines into a reduced, interpretable dimension space based on prediction similarity.

**Process**:

- **Pipeline Generation**: 20,000 ML pipelines are randomly generated. Each pipeline's configuration randomly selected from a pool of 5 imputation methods, 12 data preprocessing methods, 18 classifiers and a total of 94 hyperparameters.
- **Training & Evaluation**: These 20,000 pipelines are trained on 65 OpenML (4) datasets (binary and multi-class) using 75% of available data for a given (OpenML) dataset. They are then evaluated on the remaining 25% hold-out data.

- **Data Storage**: Hold-out data predictions from each pipeline are stored and concatenated to build a *prediction space*. Additional *meta-data* such as memory required, training time, test time, & accuracy is also stored.
- **Latent Space Creation**: Multi-dimensional scaling (MDS) is used to convert the prediction space into a 4D "configuration (latent) space" via Euclidean embedding.

**Assumptions**:

- Pipelines with similar predictions across OpenML datasets are likely to have similar predictions on unseen data.
- Pipelines with similar predictions will be closer in lower dimensional latent space.
- Proximity in the latent space implies structural or hyperparameter similarity, aiding performance understanding.

**(Phase 2): Warm Start for Bayesian Optimisation**
**Objective:** To identify optimal starting points for BO using pre-collected meta data
**Process**:
2.1 **Metadata Utilisation**: Meta data from Phase 1 is used to build a *rank order scoring mechanism*.
2.2 **Regret Measurement**: For each pipeline, *mean and median regret* were measured, i.e., how much worse a pipeline did compared to the best possible option. Lower is better. Additionally, *training time* also stored.
2.3 **Pipeline Ranking**: Pipelines are ranked for each metric (mean regret, median regret, training time). (For example, the pipeline with lowest mean regret would get rank 1 for that metric and so on.)
2.4 **Total Rank Score**: Each pipeline's ranks across all three criteria are summed to give a total score (lower is better).
2.5 **Warm Start Selection**: The 10 pipelines with the lowest total rank score are selected as warm start points for Bayesian Optimization(5).
**(Phase 3): Bayesian Optimisation sampling mechanism**
**Objective:** To efficiently explore the pipeline lower dimension configuration space for new datasets using BO
**Process**:
3.1 **New Dataset Initialisation**: When a new dataset (such as ADNI, NACC) is presented, which can comprise multimodal data if in tabular format, the warm start phase is initiated with the training subset of the new data.
3.2 **Active Sampling**: Subsequent pipeline sampling is guided by BO active sampling. BO uses a parameter kappa to control exploration vs exploitation.
3.3 **Principled constraining mechanism**:
3.3.1 The autoML system uses stored metadata (on how similar pipelines behave) to guide future choices during BO.
3.3.2 A function constraint map c estimates resource intensiveness (e.g., time, memory) each pipeline.
3.3.3 When BO selects a new pipeline, it also checks the 5 closest pipelines and picks the one with the least estimates time/memory according to c.
**(Phase 4): Ensemble generation**

**Objective:** To create a robust and accurate ensemble model or select the best single autoML pipeline
**Process**:
4.1 **Candidate Evaluation & Ranking**: Selected candidate pipelines from BO are evaluated on the training subset of the new dataset and ranked by accuracy (descending).
4.2 **Ensemble Construction**:
4.2.1 The highest ranked pipeline is added to the ensemble.
4.2.2 Its predictions are generated on the validation subset of new data.
4.2.3 Subsequent pipelines are considered:
4.2.3.1 Their predictions are compared to the current ensemble using Hamming distance.
4.2.3.2 If Hamming Distance exceeds a predefined **patience threshold** (e.g., 2%), the pipeline is discarded.
4.2.3.3 If within the threshold, the pipeline is **provisionally added**, and ensemble validation accuracy is re-evaluated.
4.2.3.4 The pipeline is **retained in the ensemble** only if it improves validation accuracy by at least a **minimum gain threshold** (e.g., 1%). Otherwise, it is discarded.
4.2.4 This process continues iteratively until:

- All pipelines have been considered, or
- No improvement is observed for **20 consecutive iterations**.

4.3 **Final Model selection & Evaluation**:

- If the final ensemble has more than one pipeline, they are combined using a **stacked ensemble**
- Otherwise, the most accurate individual pipeline is retained as the final model.

The final model (stacked or single pipeline) is evaluated on the **hold-out test set**, producing the final outcome.

To clarify the practical configuration of the AutoML search space used in this study, Table S1.X summarises the component library from which candidate pipelines were constructed, including imputation strategies, preprocessing operators, classifiers, and the number of associated tunable hyperparameters.

| Pipeline stage | Component | No. of hyperparameters | Library |
|---|---|---|---|
| **Imputation** | Mean substitution | 0 | scikit-learn |
| **Imputation** | Median substitution | 0 | scikit-learn |
| **Imputation** | Mode substitution | 0 | scikit-learn |
| **Imputation** | KNN imputation | 2 | scikit-learn |
| **Imputation** | Multivariate imputation | 2 | scikit-learn |
| **Categorical feature processing** | Ordinal Encoder | 0 | scikit-learn |
| **Categorical feature processing** | One-Hot Encoder | 0 | scikit-learn |

| | | | |
|---|---|---|---|
| **Numerical feature preprocessing** | Standard Scaling | 0 | scikit-learn |
| **Numerical feature preprocessing** | MaxAbs Scaling | 0 | scikit-learn |
| **Numerical feature preprocessing** | Robust Scaler | 0 | scikit-learn |
| **Numerical feature preprocessing** | Power Transformer | 1 | scikit-learn |
| **Numerical feature preprocessing** | Quantile Transformer | 1 | scikit-learn |
| **Numerical feature preprocessing** | Normalizer | 1 | scikit-learn |
| **Discretization** | KBins Discretizer | 3 | scikit-learn |
| **Dimensionality reduction** | FastICA | 1 | scikit-learn |
| **Dimensionality reduction** | Feature Agglomeration | 2 | scikit-learn |
| **Dimensionality reduction** | PCA | 2 | scikit-learn |
| **Dimensionality reduction** | Variance Threshold | 1 | scikit-learn |
| **Classifier** | KNN | 3 | scikit-learn |
| **Classifier** | Gaussian Process | 5 | scikit-learn |
| **Classifier** | Gaussian Naive-Bayes | 0 | scikit-learn |
| **Classifier** | Multinomial Naive-Bayes | 2 | scikit-learn |
| **Classifier** | Complement Naive-Bayes | 3 | scikit-learn |
| **Classifier** | Bernoulli Naive-Bayes | 2 | scikit-learn |
| **Classifier** | Categorical Naive-Bayes | 2 | scikit-learn |
| **Classifier** | Decision Tree | 7 | scikit-learn |
| **Classifier** | Random Forest | 7 | scikit-learn |
| **Classifier** | AdaBoost-SAMME | 3 | scikit-learn |
| **Classifier** | Gradient Boosting | 7 | scikit-learn |
| **Classifier** | Ridge | 1 | scikit-learn |
| **Classifier** | Logistic Regression | 5 | scikit-learn |
| **Classifier** | Linear SVM | 4 | scikit-learn |
| **Classifier** | Support Vector Machine | 2 | scikit-learn |
| **Classifier** | Multi-Layer Perceptron | 6 | scikit-learn |
| **Classifier** | XGBoosting | 6 | xgboost |

| | | | |
|---|---|---|---|
| **Classifier** | Relevance Vector Machine | 1 | sklearn-rvm |
| **TOTAL** | **36 components** | **82 hyperparameters** | **3 libraries** |

**Table S1.2.1**: **Pipeline component library defining the AutoML search space used in this study.** Candidate pipelines are constructed by combining imputation strategies, preprocessing operators, dimensionality-reduction methods, and classifiers drawn from the component library shown here. The table also reports the number of associated tunable hyperparameters for each component and the software library used for implementation.

*Note:* The number of hyperparameters refers to the tunable parameters associated with each pipeline component during AutoML optimisation.

#### S1.3 Evaluation Strategy

| Model | Abbreviation | Model class | Key characteristics |
|---|---|---|---|
| Logistic Regression (6) | LR | Linear | Linear decision boundary; interpretable coefficients; baseline linear classifier |
| Support Vector Machine (7) | SVM | Kernel-based | Margin-based classifier; supports non-linear decision boundaries via kernels |
| k-Nearest Neighbours (8) | KNN | Distance-based | Instance-based learning; non-parametric; local decision rules |
| Classification and Regression Tree (9) | Cart | Decision tree | Non-linear; rule-based structure; captures feature interactions |
| Random Forest (10) | RF | Ensemble (bagging) | Aggregation of decision trees; variance reduction; robust to overfitting |
| AdaBoost (11) | AdaB | Ensemble (boosting) | Sequential ensemble; focuses on difficult samples; adaptive weighting |
| Extreme Gradient Boosting (12) | XGB | Ensemble (boosting) | Gradient-boosted trees; strong non-linear modelling capacity |
| Multilayer Perceptron (13) | MLP | Neural network | Feed-forward neural network; flexible non-linear function approximation |
| Gaussian Naïve Bayes (5) | Bayes | Probabilistic | Generative classifier; assumes conditional independence |

**Table S1.3. Baseline models evaluated**

The nine individual ML models evaluated were selected to provide a heterogeneous set of linear, non-linear, probabilistic, distance-based, tree-based, ensemble, and neural network approaches. This diversity ensures coverage of different inductive biases and decision functions, enabling robust and fair comparison across modelling paradigms. All individual models were trained and evaluated using identical data splits, feature sets, and evaluation procedures.

##### S1.3.1 Stacked ensemble Model

The stacked ensemble(14) model followed a two-tier (stacked generalisation) architecture comprising multiple base learners (level-0 models) and a trainable meta-learner (level-1 model). The base learners consisted of the same nine individual machine-learning models listed in Table S1.3, spanning linear, non-linear, probabilistic, and ensemble-based approaches.

Base models were trained on the training portion of the data using internal cross-validation, and their out-of-sample predictions were used as inputs to train the meta-learner. The meta-learner was implemented as a Gaussian Naïve Bayes classifier(5), which learned how to optimally combine base-model predictions into a final decision. This approach aims to reduce variance and improve generalisation performance by leveraging complementary strengths of heterogeneous base models.

At inference time, base-model predictions for unseen test samples were first generated and then passed to the trained meta-learner to produce the final ensemble prediction. A schematic overview of the stacked ensemble architecture is provided in Supplementary Figure S1.3.

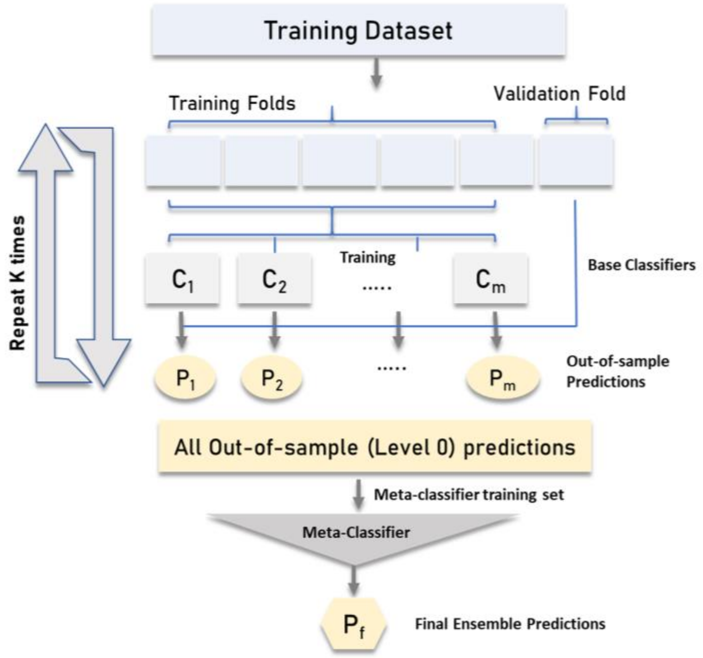


**Figure S1.3: Stacked ensemble model architecture**. A schematic illustration of the two-tier stacked ensemble model. Multiple base learners (level-0 models) are trained on the training dataset using internal cross-validation. Out-of-sample predictions from each base model are collected and used as inputs to a trainable meta-classifier (level-1 model), which produces the final prediction.

### S2. Results

#### S2.1 Per-task performance plots (mean ± SD)

Full task-level results, including mean balanced accuracy on the held-out test set and variability across 100 stratified resampling runs, are provided in the following figures. Each plot reports performance for a single prediction task and modality, with error bars indicating the standard deviation across resampling runs.

Plots are shown individually to facilitate inspection of model ranking variability and performance overlap within each clinically meaningful task.

**ADNI: sMCI vs pMCI – imaging only**

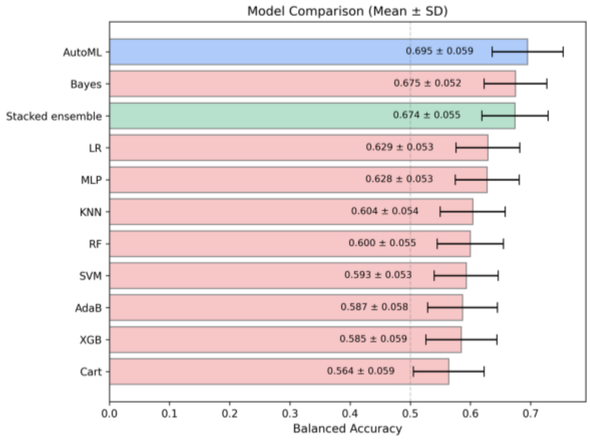


**ADNI: sMCI vs pMCI – clinical/cognitive**

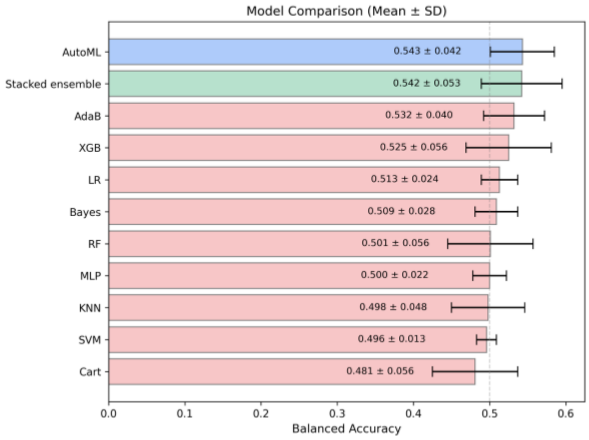


**ADNI: sMCI vs pMCI - combined**

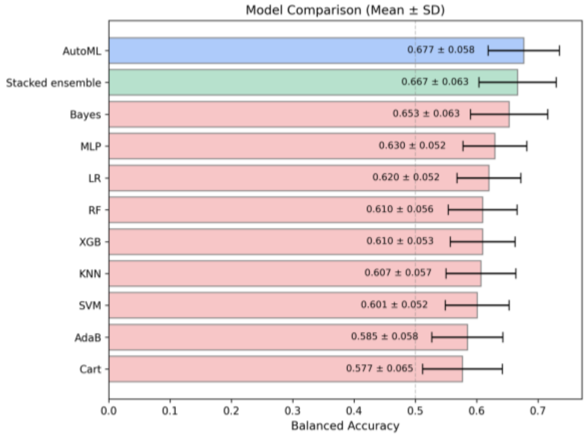


**ADNI: AD vs CN – imaging only**

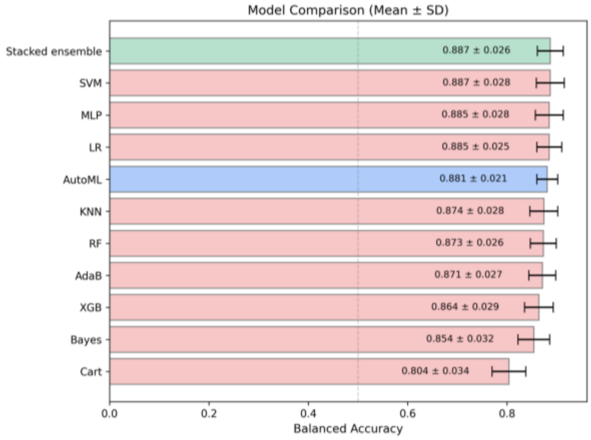


**ADNI: AD vs CN – Clinical/Cognitive**

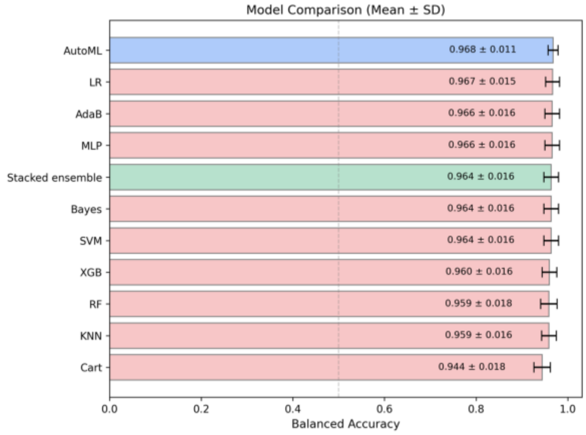


**ADNI: AD vs CN – Multimodal**

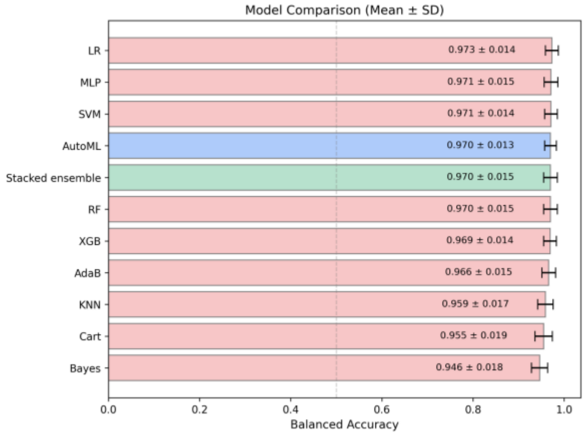

### ADNI: AD vs MCI vs CN – Clinical/Cognitive

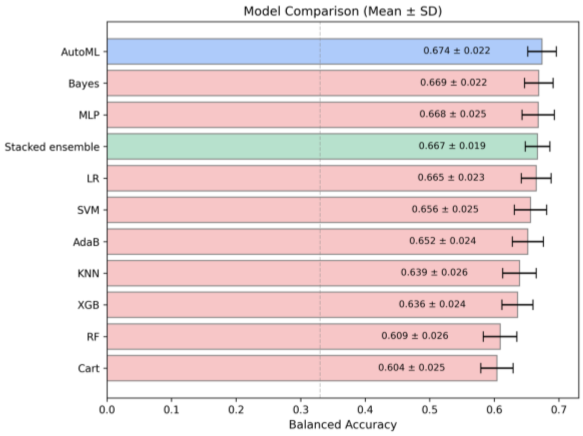


### ADNI: AD vs MCI vs CN – imaging only

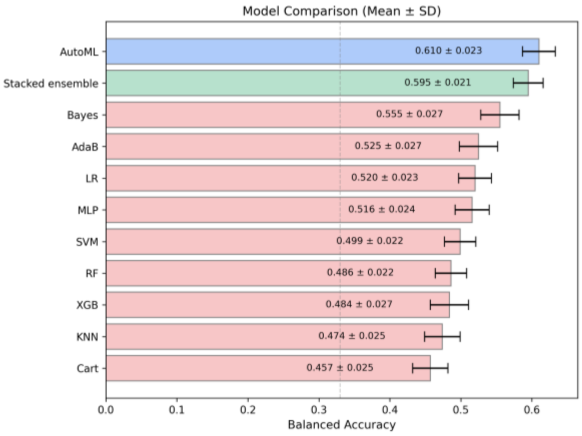


### ADNI: AD vs MCI vs CN – combined

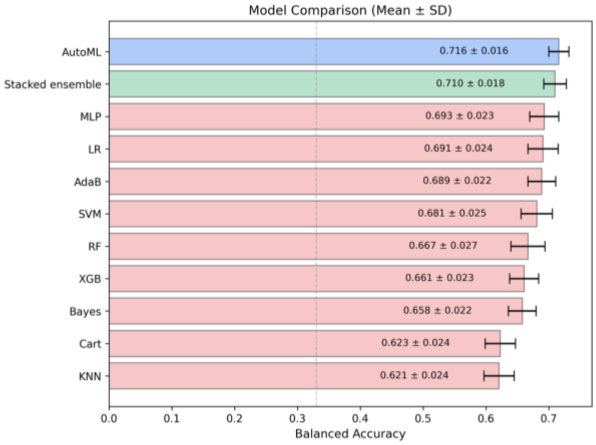


### NACC: sMCI vs pMCI – imaging only

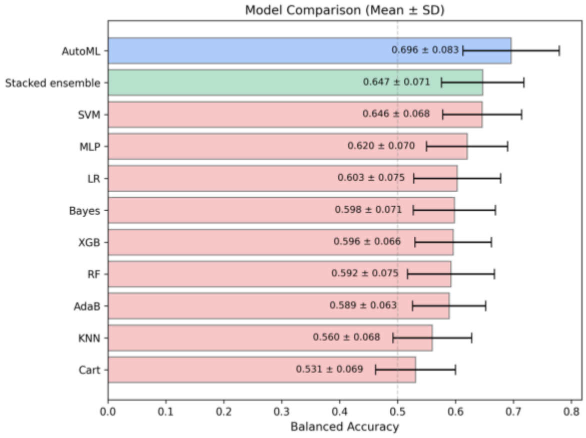


### NACC: sMCI vs pMCI – Clinical/Cognitive

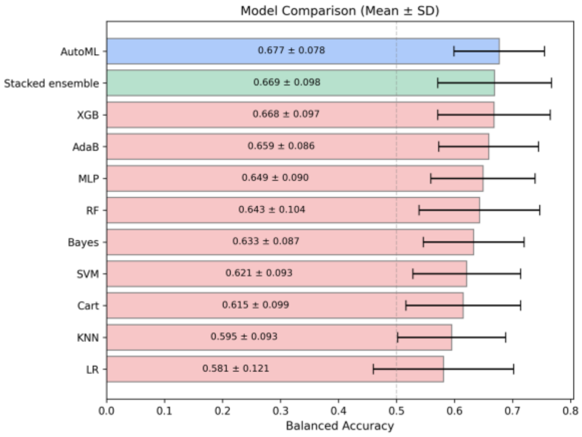


### NACC: sMCI vs pMCI – Multimodal

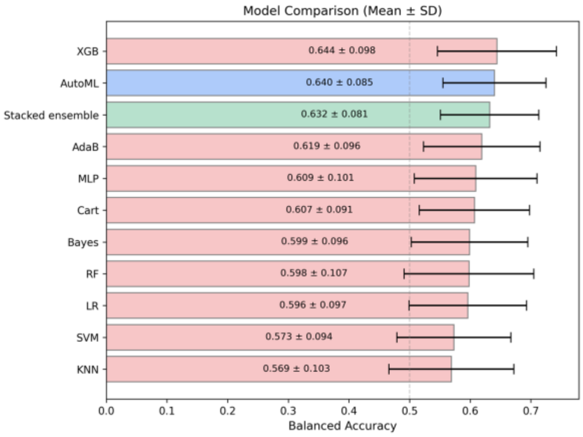

### NACC: AD vs MCI – Clinical/Cognitive

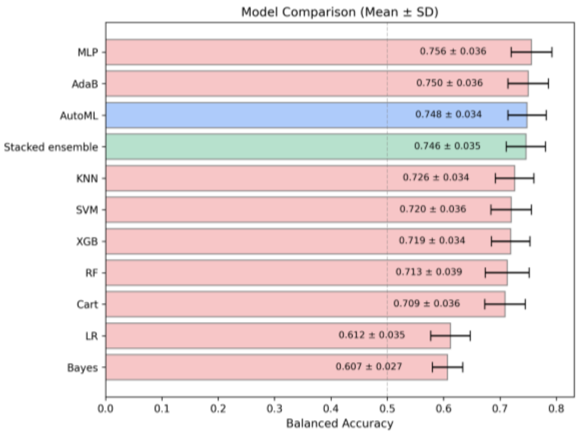


### NACC: AD vs MCI – Imaging only

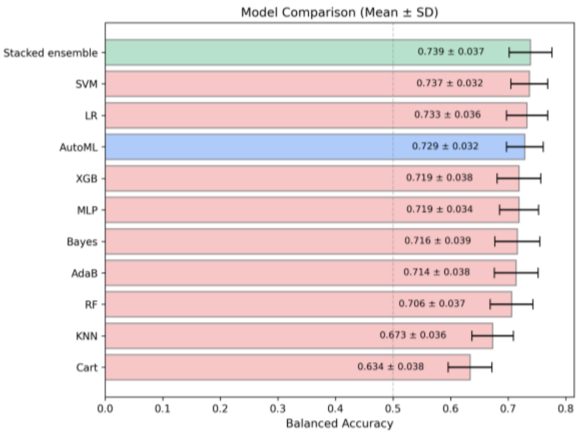


### NACC: AD vs MCI – Multimodal

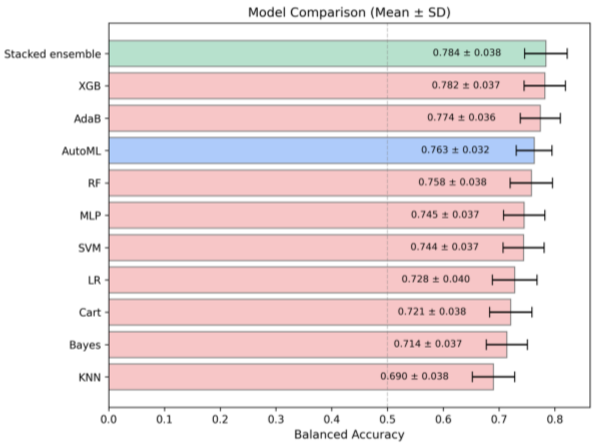


### NACC: AD vs MCI – CSF

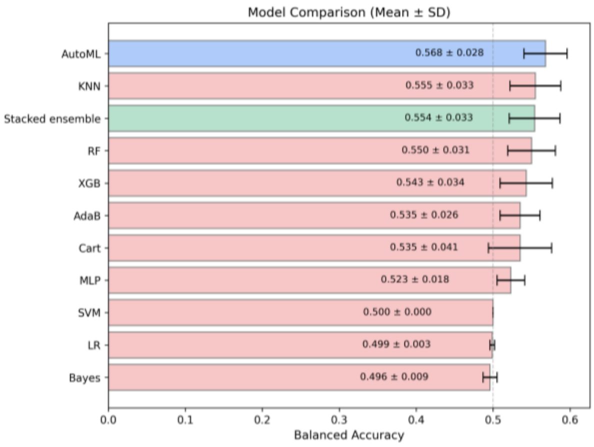


### NACC: CN vs MCI – Imaging only

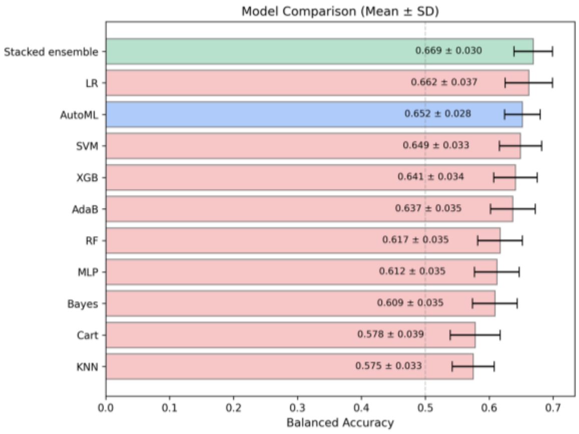


### NACC: CN vs MCI – Clinical/Cognitive

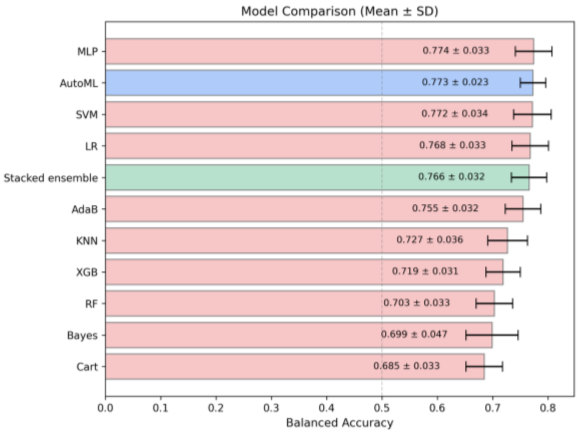

**NACC: CN vs MCI – Multimodal**

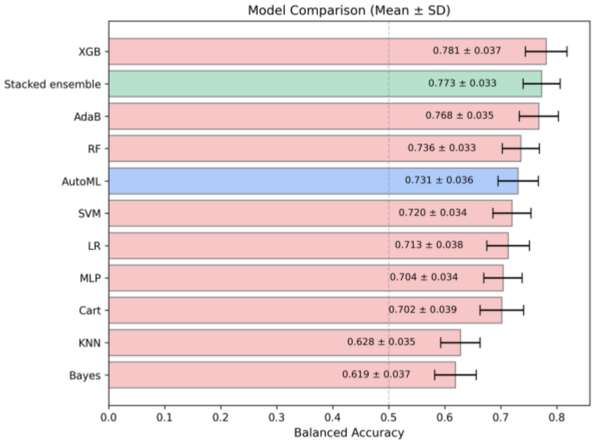


**NACC: CN vs MCI – CSF**

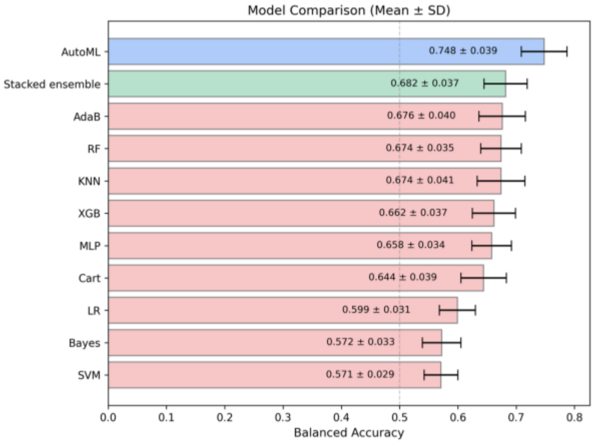

| Tasks | AutoML | Stacked | Bayes | LR | MLP | KNN | RF | SVM | AdaB | XGB | Cart |
|---|---|---|---|---|---|---|---|---|---|---|---|
| **ADNI** | | | | | | | | | | | |
| sMCI vs. pMCI – Imaging | 0.695 ± 0.059 | 0.674 ± 0.055 | 0.675 ± 0.052 | 0.629 ± 0.053 | 0.628 ± 0.053 | 0.604 ± 0.054 | 0.600 ± 0.055 | 0.593 ± 0.053 | 0.587 ± 0.058 | 0.585 ± 0.059 | 0.564 ± 0.059 |
| sMCI vs. pMCI - Clinical/Cognitive | 0.543 ± 0.042 | 0.542 ± 0.053 | 0.509 ± 0.028 | 0.513 ± 0.024 | 0.500 ± 0.022 | 0.498 ± 0.048 | 0.501 ± 0.056 | 0.496 ± 0.013 | 0.532 ± 0.040 | 0.525 ± 0.056 | 0.481 ± 0.056 |
| sMCI vs. pMCI - Multimodal | 0.677 ± 0.058 | 0.667 ± 0.063 | 0.653 ± 0.063 | 0.620 ± 0.052 | 0.630 ± 0.052 | 0.607 ± 0.057 | 0.610 ± 0.056 | 0.601 ± 0.052 | 0.585 ± 0.058 | 0.610 ± 0.053 | 0.577 ± 0.065 |
| AD vs. CN - Imaging | 0.881 ± 0.021 | 0.887 ± 0.026 | 0.854 ± 0.032 | 0.885 ± 0.025 | 0.885 ± 0.028 | 0.874 ± 0.028 | 0.873 ± 0.026 | 0.887 ± 0.028 | 0.871 ± 0.027 | 0.864 ± 0.029 | 0.804 ± 0.034 |
| AD vs. CN - Clinical/Cognitive | 0.968 ± 0.011 | 0.964 ± 0.016 | 0.964 ± 0.016 | 0.967 ± 0.015 | 0.966 ± 0.016 | 0.959 ± 0.016 | 0.959 ± 0.018 | 0.964 ± 0.016 | 0.966 ± 0.016 | 0.960 ± 0.016 | 0.944 ± 0.018 |
| AD vs. CN - Multimodal | 0.970 ± 0.013 | 0.970 ± 0.015 | 0.946 ± 0.018 | 0.973± 0.014 | 0.971 ± 0.015 | 0.959 ± 0.017 | 0.970 ± 0.015 | 0.971 ± 0.014 | 0.966 ± 0.015 | 0.969 ± 0.014 | 0.955 ± 0.019 |
| AD vs. MCI vs. CN - Imaging | 0.610 ± 0.023 | 0.595 ± 0.021 | 0.555 ± 0.027 | 0.520 ± 0.023 | 0.516 ± 0.024 | 0.474 ± 0.025 | 0.486 ± 0.022 | 0.499 ± 0.022 | 0.525 ± 0.027 | 0.484 ± 0.027 | 0.457 ± 0.025 |
| AD vs. MCI vs. CN - Clinical/Cognitive | 0.674 ± 0.022 | 0.667 ± 0.019 | 0.669 ± 0.022 | 0.665 ± 0.023 | 0.668 ± 0.025 | 0.639 ± 0.026 | 0.609 ± 0.026 | 0.656 ± 0.025 | 0.652 ± 0.024 | 0.636 ± 0.024 | 0.604 ± 0.025 |
| AD vs. MCI vs. CN - Multimodal | 0.716 ± 0.016 | 0.710 ± 0.018 | 0.658 ± 0.022 | 0.691 ± 0.024 | 0.693 ± 0.023 | 0.621 ± 0.024 | 0.667 ± 0.027 | 0.681 ± 0.025 | 0.689 ± 0.022 | 0.661 ± 0.023 | 0.623 ± 0.024 |
| **NACC** | | | | | | | | | | | |
| AD vs. MCI - Imaging | 0.729 ± 0.032 | 0.739 ± 0.037 | 0.716 ± 0.039 | 0.733 ± 0.036 | 0.719 ± 0.034 | 0.673 ± 0.036 | 0.706 ± 0.037 | 0.737 ± 0.032 | 0.714 ± 0.038 | 0.719 ± 0.038 | 0.634 ± 0.038 |
| AD vs. MCI - Clinical/Cognitive | 0.748 ± 0.034 | 0.746 ± 0.035 | 0.607 ± 0.027 | 0.612 ± 0.035 | 0.756 ± 0.036 | 0.726 ± 0.034 | 0.713 ± 0.039 | 0.720 ± 0.036 | 0.750 ± 0.036 | 0.719 ± 0.034 | 0.709 ± 0.036 |
| AD vs. MCI – Multimodal | 0.763 ± 0.032 | 0.784 ± 0.038 | 0.714 ± 0.037 | 0.728 ± 0.040 | 0.745 ± 0.037 | 0.690 ± 0.038 | 0.758 ± 0.038 | 0.744 ± 0.037 | 0.774 ± 0.036 | 0.782 ± 0.037 | 0.721 ± 0.038 |
| AD vs. MCI – CSF | 0.568 ± 0.028 | 0.554 ± 0.033 | 0.496 ± 0.009 | 0.499 ± 0.003 | 0.523 ± 0.018 | 0.555 ± 0.033 | 0.550 ± 0.031 | 0.500 ± 0.000 | 0.535 ± 0.026 | 0.543 ± 0.034 | 0.535 ± 0.041 |
| CN vs. MCI – Imaging | 0.652 ± 0.028 | 0.669 ± 0.030 | 0.609 ± 0.035 | 0.662 ± 0.037 | 0.612 ± 0.035 | 0.575 ± 0.033 | 0.617 ± 0.035 | 0.649 ± 0.033 | 0.637 ± 0.035 | 0.641 ± 0.034 | 0.578 ± 0.039 |
| CN vs. MCI - Clinical/Cognitive | 0.773 ± 0.023 | 0.766 ± 0.032 | 0.699 ± 0.047 | 0.768 ± 0.033 | 0.774 ± 0.033 | 0.727 ± 0.036 | 0.703 ± 0.033 | 0.772 ± 0.034 | 0.755 ± 0.032 | 0.719 ± 0.031 | 0.685 ± 0.033 |
| CN vs. MCI – Multimodal | 0.731 ± 0.036 | 0.773 ± 0.033 | 0.619 ± 0.037 | 0.713 ± 0.038 | 0.704 ± 0.034 | 0.628 ± 0.035 | 0.736 ± 0.033 | 0.720 ± 0.034 | 0.768 ± 0.035 | 0.781 ± 0.037 | 0.702 ± 0.039 |
| CN vs. MCI – CSF | 0.748 ± 0.039 | 0.682 ± 0.037 | 0.572 ± 0.033 | 0.599 ± 0.031 | 0.658 ± 0.034 | 0.674 ± 0.041 | 0.674 ± 0.035 | 0.571 ± 0.029 | 0.676 ± 0.040 | 0.662 ± 0.037 | 0.644 ± 0.039 |
| sMCI vs. pMCI - Imaging | 0.696 ± 0.083 | 0.647 ± 0.071 | 0.598 ± 0.071 | 0.603 ± 0.075 | 0.620 ± 0.070 | 0.560 ± 0.068 | 0.592 ± 0.075 | 0.646 ± 0.068 | 0.589 ± 0.063 | 0.596 ± 0.066 | 0.531 ± 0.069 |
| sMCI vs. pMCI - Clinical/Cognitive | 0.677 ± 0.078 | 0.669 ± 0.098 | 0.633 ± 0.087 | 0.581 ± 0.121 | 0.649 ± 0.090 | 0.595 ± 0.093 | 0.643 ± 0.104 | 0.621 ± 0.093 | 0.659 ± 0.086 | 0.668 ± 0.097 | 0.615 ± 0.099 |
| sMCI vs. pMCI – Multimodal | 0.640 ± 0.085 | 0.632 ± 0.081 | 0.599 ± 0.096 | 0.596 ± 0.097 | 0.609 ± 0.101 | 0.569 ± 0.103 | 0.598 ± 0.107 | 0.573 ± 0.094 | 0.619 ± 0.096 | 0.644 ± 0.098 | 0.607 ± 0.091 |

**Table S2.1**: **Task-level predictive performance across 20 Alzheimer's disease prediction settings in ADNI and NACC.** Mean balanced accuracy (± standard deviation across 100 resampling runs) is reported for AutoML-Multiverse, the stacked ensemble, and nine baseline machine-learning models for each prediction setting. All values are computed on unseen test sets under identical feature sets and train–test partitions within each setting.

To provide additional transparency regarding pipeline diversity, we summarise the most frequently selected AutoML-Multiverse pipeline types across tasks and modalities. Table S2.2 reports ensemble formation frequency and the top five recurrent pipeline types for each task-modality configuration.

| Task | Input Features | Ensemble formation frequency (n/100 runs) | Top 5 recurrent pipeline types (frequency across runs) |
|---|---|---|---|
| **ADNI** | | | |
| AD vs. CN | Imaging | 6 | RidgeClassifier: 27<br>SVC: 26<br>LinearSVC: 15<br>LogisticRegression: 13<br>KNeighborsClassifier: 12 |
| | Clinical/Cognitive | 0 | LogisticRegression: 31<br>RandomForestClassifier: 19<br>LinearSVC: 15<br>GaussianNB: 7<br>KNeighborsClassifier: 6 |
| | Multimodal | 0 | SVC: 32<br>LogisticRegression: 23<br>RandomForestClassifier: 21<br>LinearSVC: 3<br>GradientBoostingClassifier: 3 |
| AD vs. MCI vs. CN | Imaging | 11 | KNeighborsClassifier: 59<br>RandomForestClassifier: 16<br>SVC: 12<br>LinearSVC: 5<br>LogisticRegression: 4 |
| | Clinical/Cognitive | 54 | RandomForestClassifier: 46<br>LogisticRegression: 31<br>SVC: 23<br>KNeighborsClassifier: 22<br>LinearSVC: 10 |
| | Multimodal | 42 | LogisticRegression: 71<br>SVC: 38<br>RandomForestClassifier: 13<br>KNeighborsClassifier: 6<br>LinearSVC: 5 |
| sMCI vs. pMCI | Imaging | 42 | KNeighborsClassifier: 46<br>SVC: 27<br>LogisticRegression: 25<br>LinearSVC: 20<br>RidgeClassifier: 19 |
| | Clinical/Cognitive | 28 | BernoulliNB: 32<br>RidgeClassifier: 23<br>SVC: 17<br>LogisticRegression: 17<br>LinearSVC: 15 |
| | Multimodal | 34 | SVC: 43<br>KNeighborsClassifier: 33<br>LinearSVC: 27<br>LogisticRegression: 18<br>RandomForestClassifier: 13 |
| **NACC** | | | |
| AD vs. MCI | Imaging | 32 | SVC: 42<br>KNeighborsClassifier: 39<br>LogisticRegression: 20 |

| | | | |
|---|---|---|---|
| | | | RandomForestClassifier: 18<br>LinearSVC: 5 |
| | Clinical/Cognitive | 26 | RandomForestClassifier: 33<br>SVC: 19<br>KNeighborsClassifier: 16<br>LogisticRegression: 16<br>LinearSVC: 13 |
| | CSF | 60 | KNeighborsClassifier: 108<br>RandomForestClassifier: 26<br>SVC: 22<br>LinearSVC: 11<br>LogisticRegression: 9 |
| | Multimodal | 25 | RandomForestClassifier: 64<br>SVC: 17<br>AdaBoostClassifier: 8<br>LogisticRegression: 7<br>KNeighborsClassifier: 5 |
| CN vs. MCI | Imaging | 35 | SVC: 24<br>LogisticRegression: 22<br>RidgeClassifier: 19<br>LinearSVC: 16<br>MultinomialNB: 14 |
| | Clinical/Cognitive | 2 | LogisticRegression: 36<br>SVC: 31<br>RandomForestClassifier: 9<br>LinearSVC: 8<br>GaussianNB: 4 |
| | CSF | 8 | KNeighborsClassifier: 56<br>LogisticRegression: 19<br>SVC: 10<br>LinearSVC: 8<br>RandomForestClassifier: 4 |
| | Multimodal | 34 | SVC: 49<br>GaussianNB: 14<br>MultinomialNB: 13<br>AdaBoostClassifier: 10<br>ComplementNB: 9 |
| sMCI vs. pMCI | Imaging | 20 | GaussianNB: 34<br>BernoulliNB: 33<br>LogisticRegression: 12<br>CategoricalNB: 9<br>RandomForestClassifier: 8 |
| | Clinical/Cognitive | 32 | BernoulliNB: 42<br>GaussianNB: 33<br>CategoricalNB: 15<br>ComplementNB: 12<br>DecisionTreeClassifier: 9 |
| | Multimodal | 29 | LogisticRegression: 26<br>ComplementNB: 23<br>BernoulliNB: 17<br>RandomForestClassifier: 15<br>GaussianNB: 10 |

**Table S2.2.** Ensemble formation frequency and most recurrent AutoML-Multiverse pipeline types across task-modality configurations. Frequencies indicate the number of resampling runs (out of 100 per configuration) in which a pipeline containing the specified classifier appeared among the top five ranked pipelines.
